# Comparative Spatiotemporal Analysis of Global HIV-1 Subtype C Hotspots: Applying Bayesian Hierarchical Modeling, SaTScan, and Getis-Ord Gi* Statistics

**DOI:** 10.64898/2026.07.08.26357603

**Authors:** Qiancheng Ma, Tianzhen Zhang, Wen Yuan Zou, Liuying Lin, Siti Aisah binti Mokhtar

## Abstract

**Background:** Despite the global importance of HIV-1 subtype C, a global-scale GIS characterization of its geographic clustering and the temporal persistence of hotspots is lacking, and systematic cross-method comparisons are scarce.

**Methods:** We assembled 2,220 country–year observations from 111 countries (2005–2024), comprising 161,025 subtype C sequences, and generated internally standardized expected counts. We compared hotspot detection using Getis-Ord Gi* statistics (ArcGIS), SaTScan space–time scan statistics, and Bayesian hierarchical models with spatial–temporal smoothing, and quantified temporal persistence and cross-model concordance.

**Results:** Documented subtype C sequences showed increasing geographic concentration over time, shifting from relatively widespread detection toward progressively localized clustering, with the strongest and intensifying concentration in Southern Africa. SaTScan and Bayesian models identified fewer hotspots but showed greater temporal stability, whereas Gi* detected more localized and short-term spatial fluctuations. High-stability hotspots (sustained multi-year detection) were predominantly in Southern Africa. Zimbabwe was the only country classified as a high-stability hotspot across all three frameworks; Eswatini, Botswana, Malawi, and South Africa showed high stability in at least two models, indicating robust, model-consistent persistence.

**Conclusions:** Integrating complementary hotspot methods reveals both convergent and method-specific patterns and provides a quantitative basis to prioritize long-term persistence for targeted surveillance, resource allocation, and precision prevention.

## 1. Introduction

Acquired immunodeficiency syndrome (AIDS) remains one of the leading causes of human mortality worldwide. Human immunodeficiency virus (HIV) is widely distributed across the globe. In 2024, an estimated 1.3 million [1.0–1.7 million] new HIV infections were reported worldwide, while 630,000 [490,000–820,000] deaths were attributed to AIDS-related illnesses, corresponding to approximately one death per minute globally (UNAIDS, 2025).

HIV is transmitted through the exchange of bodily fluids. Common routes of transmission include sexual contact, sharing needles during drug use, mother-to-child transmission, accidental exposure through surgical or traumatic injuries (such as blood exchange resulting from physical violence or traffic accidents), blood transfusion, and medical-related incidents. Historically, HIV transmission was dominated by injection drug use; however, over time, sexual transmission has become the predominant route globally.

Following HIV infection, the virus progressively impairs the host immune system by destroying CD4□ T lymphocytes, leading to immunodeficiency and a wide range of secondary opportunistic infections that can ultimately result in death. At present, HIV infection is still considered incurable. Clinical management relies primarily on antiretroviral therapy (ART), which suppresses viral replication and reduces viral load, thereby prolonging the lifespan of infected individuals.

HIV exhibits substantial genetic diversity and is broadly classified into two types: HIV-1 and HIV-2. Globally, HIV-1 is the dominant type responsible for the vast majority of infections. HIV-1 is further divided into multiple subtypes, primarily subtype B and non-B subtypes. Subtype B, being predominant in Western countries, has been extensively studied over the past several decades. However, non-B subtypes account for the majority of global HIV-1 infections, representing approximately 77% of cases worldwide (Tong et al., 2005).

Among non-B subtypes, subtype C is the most prevalent, accounting for approximately 46.6% of all HIV-1 infections globally (Hemelaar et al., 2019). Different HIV subtypes often exhibit substantial differences in drug resistance profiles, vaccine development challenges, diagnostic performance, and transmission dynamics (Lessells et al., 2012; Ng’uni et al., 2020; Apetrei et al., 1996; Taylor et al., 2008). Understanding the geographic distribution of HIV subtypes is therefore critical for guiding national and international public health agencies, such as Centers for Disease Control, in allocating resources and prioritizing research investments.

At present, international organizations such as the World Health Organization (WHO) and UNAIDS lack comprehensive databases reporting country–year–subtype–specific HIV counts. Consequently, researchers seeking to examine the distribution of HIV subtypes across countries must rely primarily on data released by national public health agencies and on integrated sequence-sharing platforms led by the National Center for Biotechnology Information (NCBI), the DNA Data Bank of Japan (DDBJ), and the European Nucleotide Archive (ENA) (They synchronize information across all platforms on a daily basis). These platforms accept viral genetic sequence submissions from researchers worldwide and provide invaluable resources for characterizing global HIV genetic diversity and subtype distributions (Sayers et al., 2025). HIV sequences deposited in GenBank are downloaded biweekly by the Los Alamos National Laboratory (LANL) HIV Sequence Database and undergo curation, annotation, standardization, validation, and quality control as they are integrated into the platform (Kuiken et al., 2003; National Institute of Allergy and Infectious Diseases [NIAID], 2025).

To provide an overview of the global distribution, we aggregated all subtype C sequences from the LANL database by country across the 20-year study period (2005-2025). Figure 1 presents the cumulative distribution of subtype C sequences.

**Figure 1.**
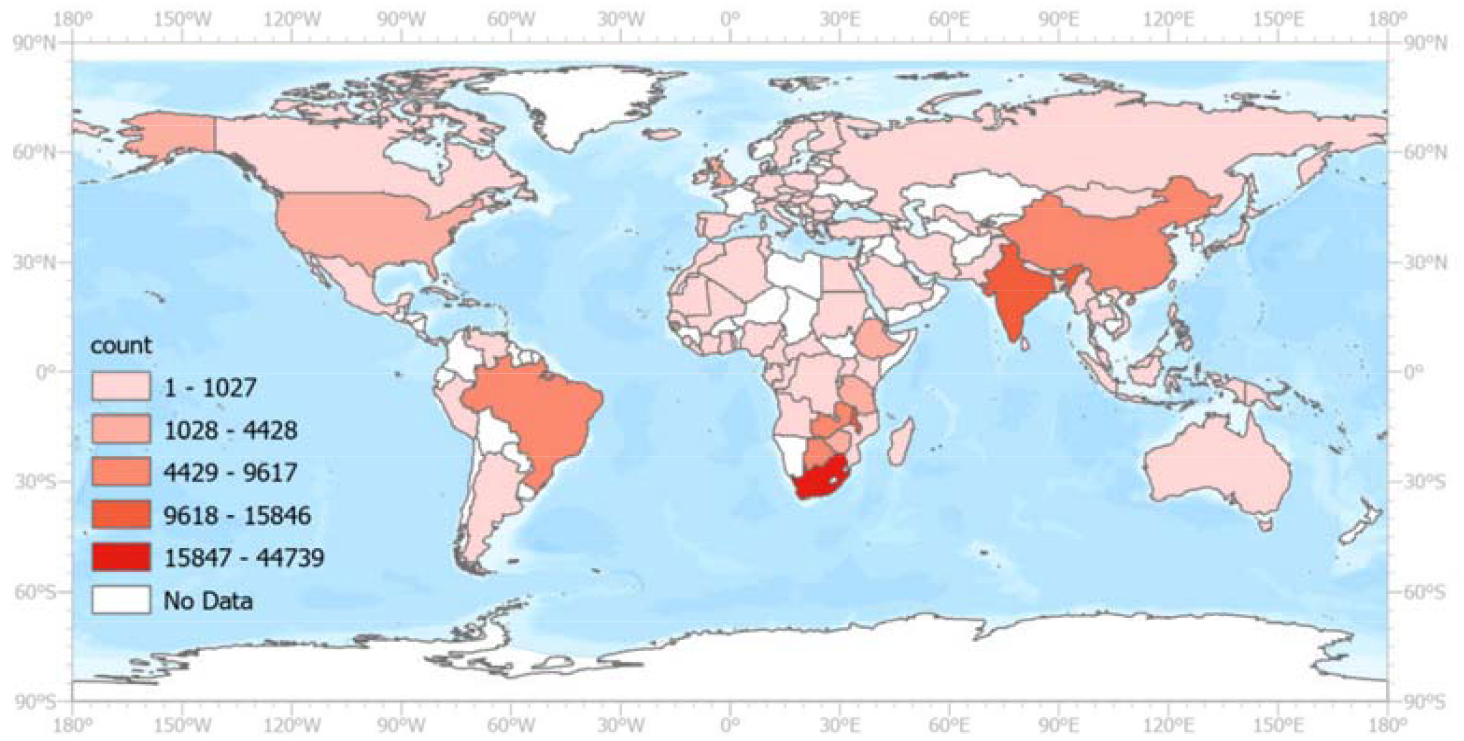
Global Distribution of Map of HIV-1 Subtype C Case.

### 1.1. Literature review

While HIV-1 subtype C has been extensively investigated, most existing studies remain confined to regional or country-specific contexts. Numerous analyses have focused on the origin, dissemination, and evolutionary dynamics of subtype C within individual settings, including South Africa, Brazil, India, and parts of Europe, primarily using molecular epidemiology and phylogenetic or phylogeographic approaches (Wilkinson et al., 2016; Delatorre et al., 2013; Bello et al., 2012; Shen et al., 2011; Alexiev et al., 2022). These studies provide important insights into local transmission histories but are inherently limited in spatial scale.

At the global level, research on HIV-1 subtype C has largely emphasized descriptive distribution patterns, evolutionary emergence, and broad circulation dynamics across regions, without applying formal spatial statistical frameworks (Li et al., 2024). Although phylodynamic and phylogeographic analyses have substantially advanced understanding of subtype C evolution and migration pathways, they do not explicitly identify geographic transmission hotspots using spatial clustering statistics.

To date, no study has conducted a global-scale spatiotemporal analysis of HIV-1 subtype C using formal spatial statistical methods, systematically compared different hotspot detection approaches, or evaluated the temporal stability of identified hotspots across multiple time periods.

A global-scale perspective is critical for understanding the geographic distribution of HIV-1 subtype C, as its transmission is fundamentally driven by cross-border and cross-regional processes rather than isolated local epidemics. The spatial burden of subtype C reflects sustained international dissemination shaped by population mobility, migration networks, and long-term diffusion across national boundaries. Consequently, analyses restricted to single countries or regions are insufficient to capture the interconnected nature of subtype C transmission. A global analytical framework is therefore necessary to identify transnational hotspot structures and to reveal spatial patterns that directly inform coordinated, cross-border prevention and control strategies.

Moreover, hotspot detection is inherently sensitive to methodological assumptions, and results derived from a single spatial approach may reflect model-specific biases rather than robust epidemiological signals. By systematically comparing multiple spatial detection methods, this study enables the identification of consistent hotspots that are stable across analytical frameworks, thereby strengthening inferential reliability. Regions detected as hotspots by multiple methods are more likely to represent persistent transmission foci, whereas method-specific signals can be interpreted with appropriate caution. This comparative strategy provides a more nuanced and credible assessment of global HIV-1 subtype C spatial risk.

Incorporating a temporal dimension further allows the distinction between transient clusters and temporally stable hotspots, which is critical for understanding long-term transmission patterns and informing sustainable public health interventions.

## 2. Materials and Methods

### 2.1. Description of the study area

As of 2025, there are 195 United Nations–recognized sovereign states, along with two UN observer states (the Holy See and the State of Palestine), and ten entities that are unrecognized or partially recognized (Taiwan, Kosovo, South Ossetia, Abkhazia, Northern Cyprus, the Sahrawi Arab Democratic Republic, Transnistria, and Somaliland), yielding a total of 205 political entities worldwide. Data included in this study cover 111 political entities (United Nations General Assembly, n.d.).

Existing descriptive studies suggest that HIV-1 subtype C is primarily distributed in Southern Africa, Eastern Africa, India, as well as in parts of southwestern China and southern Brazil (Bbosa et al., 2019; Giovanetti et al., 2020; Chen et al., 2019; Hemelaar et al., 2019).

### 2.2. Data sources

Data used in this study were obtained from the Los Alamos National Laboratory (LANL) HIV Sequence Database, which contains nearly all publicly available HIV-1 subtype C sequence data worldwide. The data acquisition mechanism of LANL operates as follows. Sequence data are primarily sourced from GenBank (NCBI). On a biweekly basis, LANL experts and staff curate, review, and update GenBank records using a combination of expert manual inspection and automated quality-control procedures before incorporating them into the LANL database (Kuiken et al., 2003; National Institute of Allergy and Infectious Diseases [NIAID], 2025).

GenBank operates within the International Nucleotide Sequence Database Collaboration (INSDC), together with the DNA Data Bank of Japan (DDBJ) and the European Nucleotide Archive (ENA). Under this framework, the three databases synchronize sequence records on a daily basis, ensuring consistency and completeness across platforms (Nakamura et al., 2013). In addition, GenBank maintains a data synchronization mechanism with GenBase, which is operated by the China National Center for Bioinformation (CNCB) (Bao & Xue, 2023).

Furthermore, CNCB collaborates with the National Primate Research Center of Thailand, Chulalongkorn University (CU-NPRC; Thailand), the Vavilov Institute of General Genetics, Russian Academy of Sciences (VIGG; Russia), King Abdullah University of Science and Technology (KAUST; Saudi Arabia), and Quaid-i-Azam University (QAU; Pakistan), together with an additional 30 organizations from 12 countries, under the Open Biodiversity and Health Big Data Alliance (BHBD). Within this framework, participating institutions promote data sharing and interoperability across platforms (Open Biodiversity and Health Big Data Alliance, n.d.).

As a result of these interconnected data-sharing and curation mechanisms, the LANL HIV Sequence Database has access to a substantial proportion of global HIV-1 subtype C sequence resources. Given the absence of dedicated sequence repositories maintained by WHO or UNAIDS, and the presence of LANL’s independent expert curation procedures, the LANL platform represents a publicly accessible database with robust quality control that captures the majority of available global HIV-1 subtype C sequence data. The detailed data sources and integration mechanisms are illustrated in Figure 2 Schematic overview of global data exchange and curation for HIV-1 subtype C sequences

**Figure 2.**
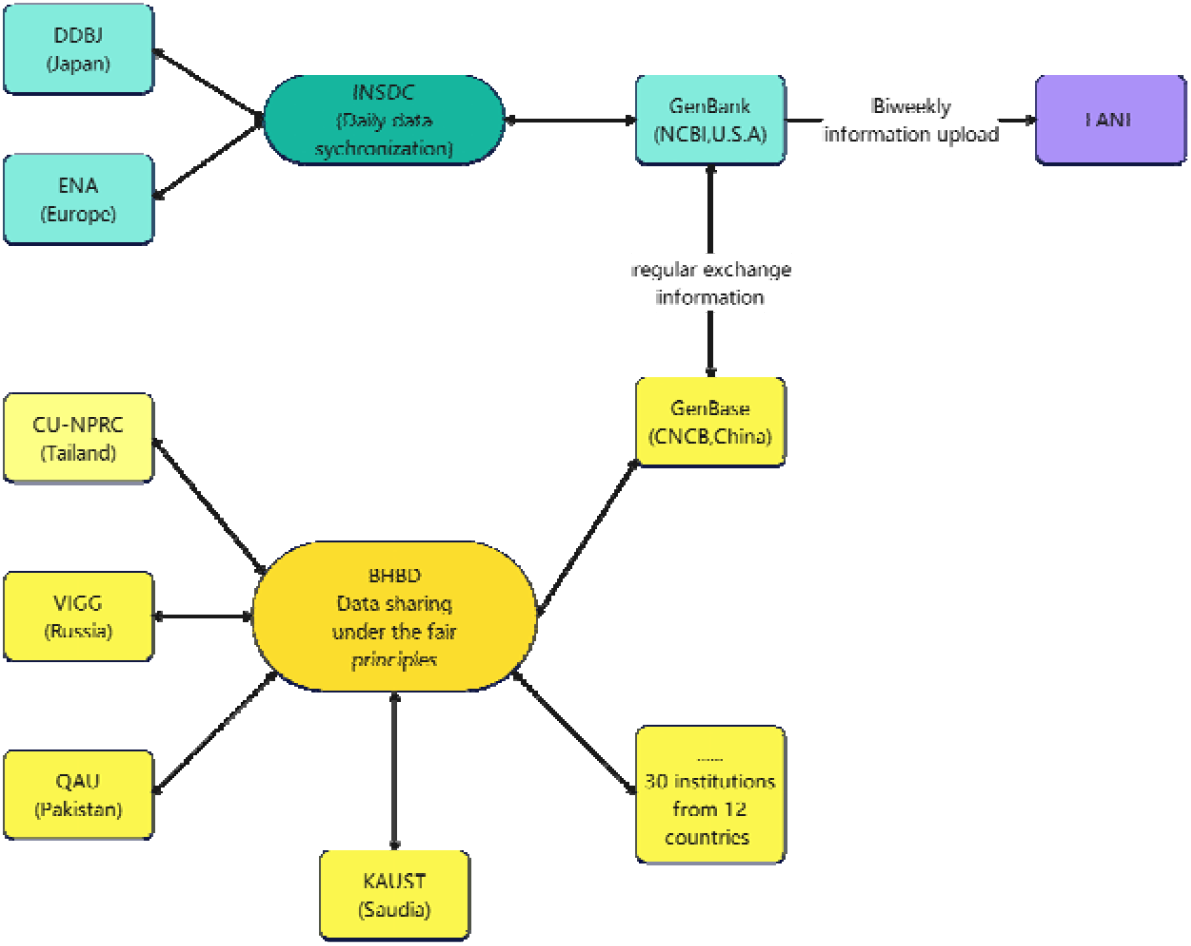
Schematic overview of global data exchange and curation for HIV-1 subtype C sequences.

**Figure 3.**
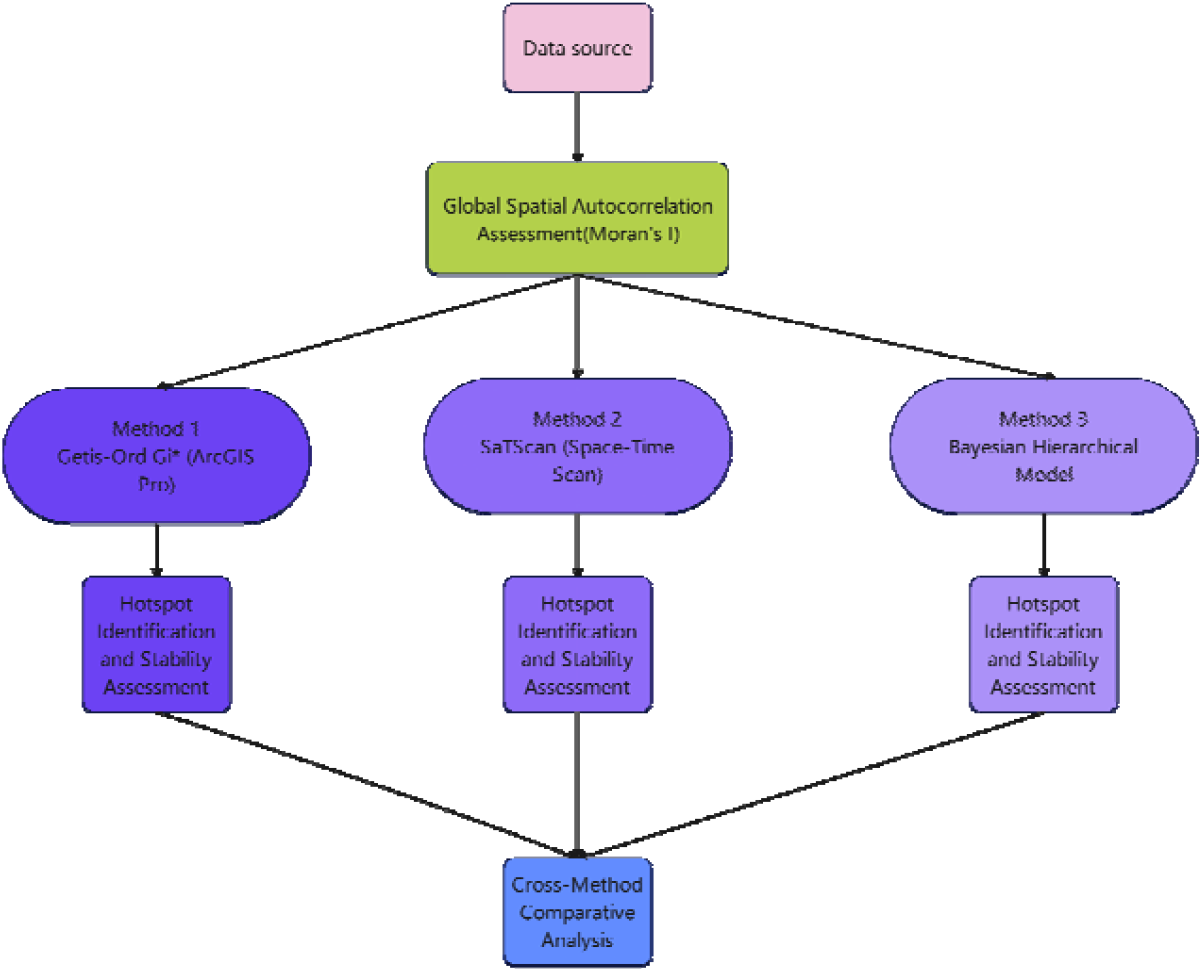
Overall analytical workflow for hotspot identification, stability assessment, and cross-method comparison.

HIV-1 subtype C data were extracted from the Los Alamos National Laboratory (LANL) HIV Sequence Database using the Sequence Search Interface, selecting subtype C and sampling years 2005–2025, with no additional restrictions applied beyond subtype and sampling year. A total of 161,098 sequences were retrieved, and metadata (including sequence ID, country, accession number, and sampling year) were saved using the Background Information function. Subsequently, the metadata were processed using Python to aggregate sequences into annual country-level case counts. Sequence counts were used as a proxy measure to characterize the spatial and temporal distribution of HIV-1 subtype C, rather than as direct estimates of clinical incidence.

Annual population estimates were obtained from the World Bank. Incidence rates per 100,000 population were calculated for each year (2005–2024) and appended as separate fields to the country attribute table.

Following data extraction and harmonization, the final country–year dataset served as a shared input for the overall workflow shown in Figure X. We first conducted a global spatial autocorrelation check (Moran’s I) to characterize whether the country-level distribution exhibited non-random spatial clustering. The dataset was then analyzed in parallel using three hotspot detection frameworks: (i) Getis–Ord Gi* in ArcGIS Pro to identify statistically significant local clusters of high values, (ii) SaTScan space–time scanning to detect significant spatiotemporal clusters, and (iii) a Bayesian hierarchical model to estimate smoothed country–year relative risks and infer hotspot states probabilistically. Within each framework, outputs were transformed into two derived products: a country–year hotspot label (hotspot vs non-hotspot) and a corresponding stability summary that captured whether hotspot signals persisted over time (i.e., “stable points” /persistent hotspots). Notably, the Bayesian module used two stability representations: a model-native stability measure based on posterior hotspot probabilities (used to summarize persistence within the Bayesian results), and a separate harmonized stability index used for cross-method evaluation. Specifically, because Gi*, SaTScan, and Bayesian models produce outputs on different statistical scales, cross-method comparison was performed after converting each method’s output into a common country–year hotspot indicator and then computing a unified stability measure on that harmonized indicator. This design allows the Bayesian model to retain its probabilistic, within-model stability characterization while still enabling like-for-like comparison of hotspot persistence across all three methods.

### 2.3. Methodology

#### 2.3.1. Spatial units and preprocessing

National administrative boundaries were obtained from the Natural Earth Admin 0 – Countries dataset and used as the spatial analysis units. All spatial layers were projected to a Cylindrical Equal Area (World) coordinate system to support global-scale comparability. Non-spatial tables (annual HIV-1 subtype C case counts and population data) were linked to country polygons using standardized ISO-A3 codes to ensure consistent joining across datasets.

#### 2.3.2. Construction of country–year case counts and incidence rates

HIV-1 subtype C sequence records were extracted from the LANL HIV Sequence Database using the Sequence Search Interface by selecting subtype C and sampling years 2005–2025, with no additional restrictions beyond subtype and sampling year. A total of 161,098 sequences were retrieved. Metadata (sequence ID, country, accession number, sampling year) were exported using the Background Information function and processed in Python to aggregate sequences into annual country-level case counts (country–year counts).

Subtype assignments in LANL are generated through a multi-tiered curation workflow that combines author-provided subtype labels, automated verification (e.g., RIP/BLAST/phylogenetic checks), and staff review for discrepant records. Given these established curation procedures, no additional manual cleaning or subtype reclassification was performed in this study.

Annual national population estimates were obtained from the World Bank. For each year (2005–2024), incidence rates per 100,000 population were computed as:

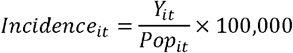

where

*Y*_*it*_ is the country–year subtype C sequence count and

*Po*_*it*_ is the corresponding annual population.

#### 2.3.3. Global spatial autocorrelation (justification for local hotspot statistics)

Before local hotspot detection, global spatial autocorrelation was assessed using Moran’s I for each year’s incidence surface to evaluate whether the spatial distribution deviated from randomness. The presence of positive spatial autocorrelation supported the use of local indicators of spatial association (LISA), particularly Getis–Ord Gi*, for detecting local clustering patterns.

#### 2.3.4. Local hotspot detection using Getis–Ord Gi* (ArcGIS Pro)

##### (a) Getis–Ord Gi* statistic

Local hotspots and coldspots were identified using the Getis–Ord Gi* statistic, which evaluates whether high (or low) values cluster spatially around each feature relative to the global mean. For a location *i*, the Gi* statistic can be expressed as:

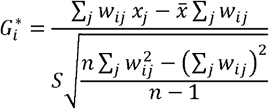

(Getis & Ord, 1992; Environmental Systems Research Institute [ESRI], 2024)

where *x*_*j*_ is the attribute value at location *j, w*_*ij*_ denotes the spatial weight between *i* and *j, n* is the number of spatial units, 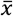 is the global mean, and *S* is the global standard deviation. ArcGIS Pro reports Gi* results as z-scores and p-values, where the z-score and p-value indicate whether the observed spatial clustering of high/low values is statistically significant under the null of spatial randomness (Esri, n.d.).

#### (b) Implementation settings in ArcGIS

Hotspot analysis was conducted using ArcGIS Pro 3.2.5 (Hot Spot Analysis: Getis–Ord Gi*). For each year, the annual incidence field (HIV_Rate_per_100k) was used as the input analysis variable. Spatial relationships were defined using a Fixed Distance Band, while the threshold distance was not manually specified; ArcGIS automatically determined an optimal neighborhood distance based on feature spatial distribution. Euclidean distance was used. To ensure comparability across years, both Self-Potential and False Discovery Rate (FDR) correction were disabled. The Gi* tool outputs, for each country-year layer, the GiZScore,*p*-value, and the ArcGIS confidence category (Gi_Bin).

##### (c) Hotspot significance classification (Gi_Bin)

ArcGIS Gi_Bin values were used to classify significance levels. Statistically significant hotspots were defined as Gi_Bin ≥ +2 (95% confidence; *p*<0.05), with Gi_Bin = +3 indicating 99% confidence hotspots. Coldspots were defined analogously (Gi_Bin ≤ −2).

##### (d) Hotspot persistence / stability index (Gi*)

To assess temporal stability of Gi*-identified hotspots, yearly Gi_Bin outputs were binarized as:

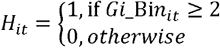

A country-level hotspot stability index over the 20-year period (2005–2024) was computed as:

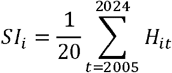

Values closer to 1 indicate highly persistent hotspots, while values approaching 0 indicate unstable or sporadic hotspot behavior.

#### 2.3.5. Spatiotemporal cluster detection using SaTScan (space–time scan statistic)

Kulldorff’s space–time scan statistic was implemented in SaTScan (v10.3.3) to identify statistically significant spatiotemporal clusters under the discrete Poisson model. The procedure scans a set of cylindrical windows across space and time, where the circular base represents the spatial footprint and the cylinder height represents the temporal interval. For each candidate cylinder *Z*, the observed number of cases inside the window*C*(*Z*) is compared with the expected number of cases*E*(*Z*) under the null hypothesis of constant risk (Kulldorff, 1997/202).

For each candidate window *Z*, SaTScan evaluates evidence of elevated risk inside the cylinder using the log-likelihood ratio (LLR). Let *C* denote the total number of cases in the entire study region and period, *C*(*Z*) the observed cases inside *Z*, and *E*(*Z*) the expected cases inside *Z* under the null. The Poisson-based LLR is computed as: (Kulldorff, 1997/2022).

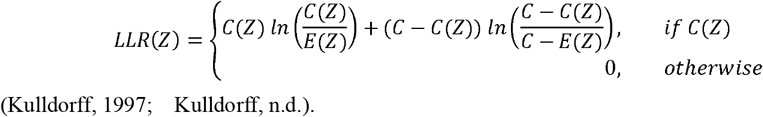

The scanning window with the maximum LLR is reported as the most likely cluster, and additional non-overlapping windows may be reported as secondary clusters. Statistical significance is assessed through Monte Carlo hypothesis testing as implemented in SaTScan, where the distribution of the maximum LLR under the null is approximated by repeated randomization; clusters with *p* <0.05 are considered statistically significant. (Kulldorff, 2022).

In this study, we set the maximum spatial cluster size to 10% of the population at risk and the maximum temporal cluster size to 50% of the study period. Statistical significance was evaluated using 999 Monte Carlo replications, and significant clusters were defined at the 5% level (*p* <0.05). (Kulldorff, 2022).

Hotspot definition (SaTScan): a country-year was labeled as hotspot if it belonged to a statistically significant high-rate space–time cluster (Monte Carlo p < 0.05).

##### (a) Hotspot significance classification (SaTScan)

SaTScan identifies statistically significant space–time clusters (cylindrical windows) rather than country–year units. In this study, a country–year was classified as a SaTScan hotspot if it fell within at least one statistically significant high-rate space–time cluster detected under the discrete Poisson model (p < 0.05). If a country–year was covered by multiple significant clusters (most likely or secondary), it was coded as a hotspot once (binary).

##### (a) Hotspot persistence / stability index (SaTScan)

To assess the temporal stability of SaTScan-identified hotspots, SaTScan cluster outputs were converted into a binary country–year hotspot matrix 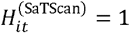 if country *i* was classified as a hotspot in year *t*, and otherwise. A country-level SaTScan hotspot stability index was then computed as:

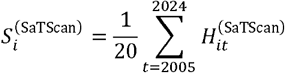

Here, 20 denotes the total number of study years (2005–2024). Values of 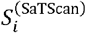 closer to 1 indicate highly persistent (“stable”) hotspots, whereas values approaching 0 indicate sporadic hotspot detection.

#### 2.3.6. Bayesian hierarchical spatiotemporal modeling (BYM2 + RW1 + interaction)

##### (b) Likelihood and relative risk formulation

A fully Bayesian hierarchical space–time model was constructed to estimate country–year relative risk (RR) while borrowing strength across space and time. Let *Y*_*it*_ denote the observed subtype C count for country *i* in year *t*. We assumed:

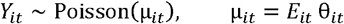

(Clayton & Kaldor, 1987)
where *E*_*it*_ is the expected count and θ_*it*_ is the relative risk (RR). Expected counts were derived using year-specific internal standardization, allocating the annual total cases proportionally to population such that∑__*i*__ *E*__*it*__ =∑__i__ *Y*_*it*_ for each year *t*.

The log-RR was modeled as:

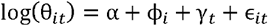

(Knorr-Held, 2000)

Where α is a global intercept, ϕ_*i*_is the spatial effect (BYM2), γ__*t*__ is the temporal effect (RW1), and ϵ_*it*_ is an IID space–time interaction term.

##### (b) Spatial component: BYM2 parameterization

The spatial effect ϕ_*i*_ was modeled using a BYM2-type decomposition that mixes a structured ICAR component and an unstructured IID component:

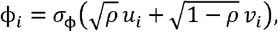

(Riebler et al., 2016)

where *u*_*i*_ is the structured spatial effect assigned an ICAR prior defined through a GMRF precision matrix*Q, v*_*i*_ ~ *N*(0, 1) is the unstructured spatial effect (IID Gaussian), *σ*_ϕ_ controls overall spatial variability, and*ρ* ∈ (0, 1)controls the mixing between structured and unstructured components. Sum-to-zero constraints were imposed for stability and identifiability by centering the latent components. In our implementation, ICAR graph scaling as in the strict BYM2 formulation was not applied; therefore, the marginal variance of the structured component depends on the specified adjacency graph.

<H5>Spatial adjacency (graph) construction

Because global country polygons can yield unstable contiguity relationships (e.g., islands), spatial adjacency was constructed using a K-nearest-neighbor (KNN) graph based on great-circle distance. Country centroid coordinates (latitude/longitude) were used to compute pairwise distances via a haversine distance matrix; each country was connected to its *k*= 5 nearest neighbors, and the adjacency matrix was used symmetrized. The resulting binary adjacency matrix (denoted*W*) was stored as a sparse matrix and to construct the GMRF precision matrix *Q* for efficient ICAR computations.

##### Temporal component: first-order random walk (RW1)

Temporal dependence was modeled using a first-order Gaussian random walk:

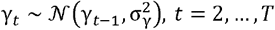

(Gelman et al., 2013)

implemented via Gaussian Random Walk with σ_γ_ HalfNormal(1.0).

##### (d) Space–time interaction (IID)

Residual space–time variation not captured by the main spatial and temporal components was modeled as an IID Gaussian interaction: 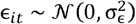. A comparatively tighter prior σ_ϵ_ ~ HalfNormal(0.3) was used to reduce overfitting and improve numerical stability, and the interaction matrix was globally centered to prevent absorption of main-effect signal.

##### (e) Priors and computation

Weakly informative priors were applied, including α ~ *N* (0,1), σ_ϕ_ ~ HalfNormal(1.0), ρ ~ Beta(2,2), σ_γ_ ~ HalfNormal(1.0), and σ_ϵ_ ~ HalfNormal(0.3), consistent with the implemented PyMC model configuration. Model inference was performed in PyMC (v5) using MCMC with 32 parallel chains, 1,000 draws and 1,000 tuning steps per chain, and a target acceptance rate of 0.95 to reduce divergent transitions.

##### (f) Bayesian hotspot classification

Country–year observations were classified as hotspots if the posterior lower bound of the 95% credible interval for RR exceeded 1 (and as coldspots if the posterior upper bound was below 1).

##### (d) Hotspot persistence / stability index (Bayesian)

In the hotspot persistence analysis, long-term stability was quantified using the posterior exceedance probability that the country–year relative risk exceeds a predefined hotspot threshold. For each country *i* and year *t*, we computed: *p*_*it*_ = *P*(*RR*_*it*_ > 1.5 | *data*) For each country *i* that was identified as a hotspot in at least one year, we then calculated the 20-year average posterior exceedance probability:

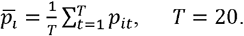

Hotspot persistence was subsequently classified based on 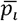 into four categories: extremely persistent 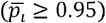, moderately persistent 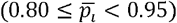, weakly persistent 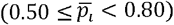, and non-persistent 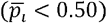.

#### 2.3.7. Cross-method comparability and hotspot stability (how you “systematically compare”)

To enable method comparison on a common scale, outputs from Gi*, SaTScan, and Bayesian modeling were harmonized into a country–year binary hotspot indicator (1 = hotspot; 0 = non-hotspot) using the method-specific rules described above (Gi_Bin ≥ 2; membership in any significant SaTScan cluster; Bayesian RR credible-interval rule). A unified stability index was then defined as the mean hotspot indicator over the 20-year period (2005–2024), and categorized into high/medium/low stability using predefined thresholds (≥0.6; 0.5–0.6; <0.5).

## 3. Result

### 3.1. Overall data overview and temporal pattern of subtype C sequence availability (2005–2024)

The dataset consisted of 2,220 country–year records from 111 countries spanning 2005–2024, with a total of 161,025 HIV-1 subtype C sequences. Expected case counts generated through internal standardization closely matched the observed totals (ΣO = 161,025; ΣE = 161,040; ratio = 1.0001), confirming internal numerical consistency. During preprocessing, a small number of records were excluded because LANL location labels could not be mapped one-to-one to ISO A3 country codes, meaning that a small subset of political entities could not be incorporated into the country–year panel. In addition, 2025 records were excluded because only 10 sequences were available for that year, providing insufficient information for meaningful annual inference and potentially introducing instability in temporal comparisons.

Figure 4 summarizes the annual number of subtype C sequences captured by LANL. Between 2005 and 2008, yearly totals were approximately 10,000–14,000 sequences, followed by a decline during 2009–2012. Submissions increased again from 2013 and peaked during 2017–2019 (approximately 16,000–17,000 sequences per year). After 2019, the number of newly captured sequences decreased sharply. Importantly, because LANL sequence counts reflect the timing and intensity of sequencing, submission, and curation workflows (rather than population-based surveillance), inter-annual fluctuations should be interpreted primarily as changes in data deposition and availability, not as direct estimates of global HIV incidence.

**Figure 4.**
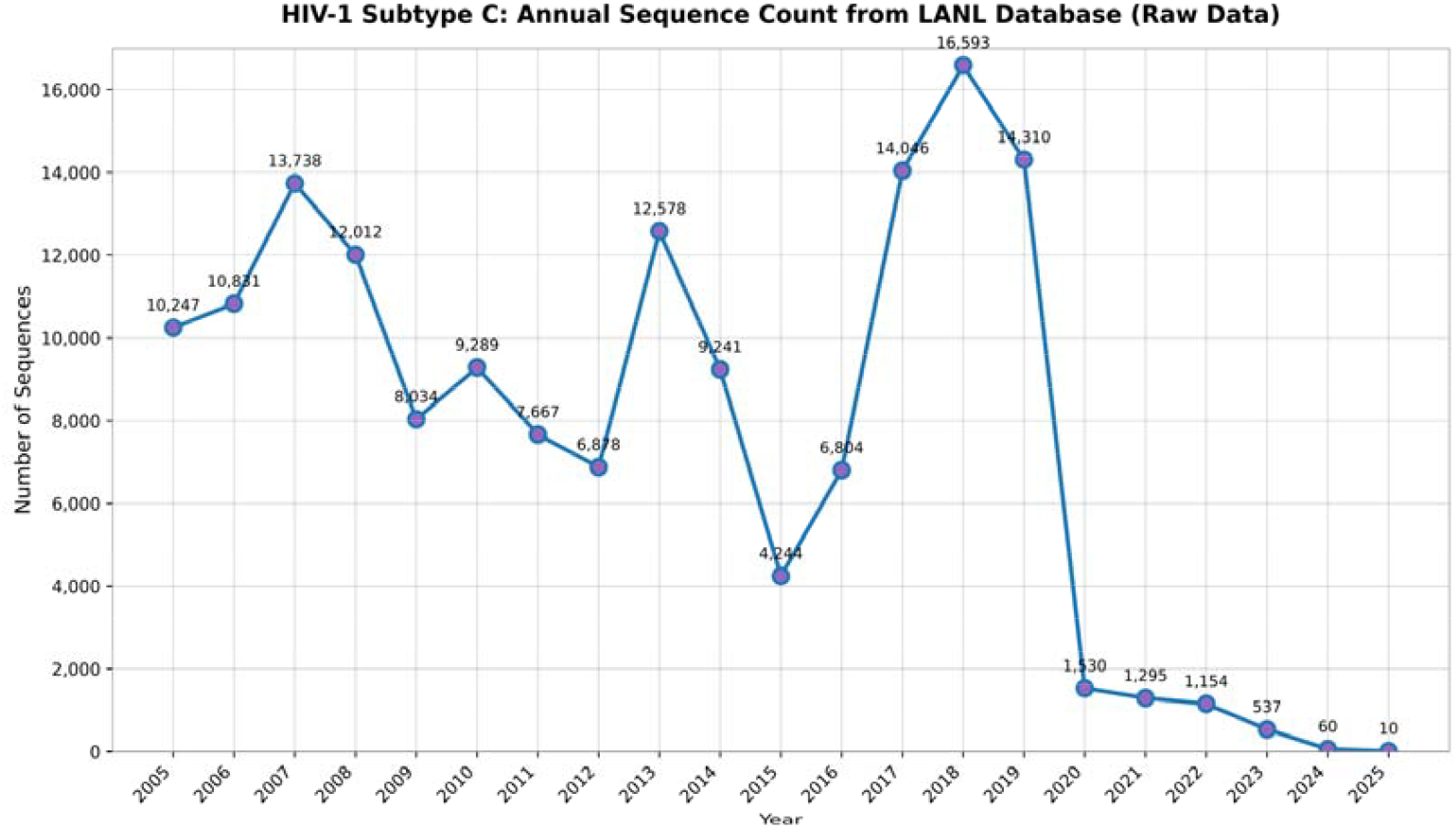
Annual global count of HIV-1 subtype C sequences, 2005–2024.

To contextualize the post-2019 decline in sequence counts, Figure 5 presents the global annual number of new HIV infections (UNAIDS). Global new infections have declined substantially over the past decades, reflecting long-term progress in prevention and treatment scale-up. In addition, the COVID-19 pandemic caused widespread disruption to essential health services, including HIV testing and diagnosis pathways, which plausibly reduced opportunities for case detection and subsequent sequencing/sequence deposition during and after 2020. Therefore, the marked reduction in LANL sequence capture after 2019 is most plausibly explained by a combination of (i) pandemic-related service and laboratory disruptions affecting testing and downstream sequencing/submission workflows, and (ii) the broader long-term decline in global new HIV infections, rather than indicating that the LANL resource was “abandoned” or no longer relevant.

**Figure 5.**
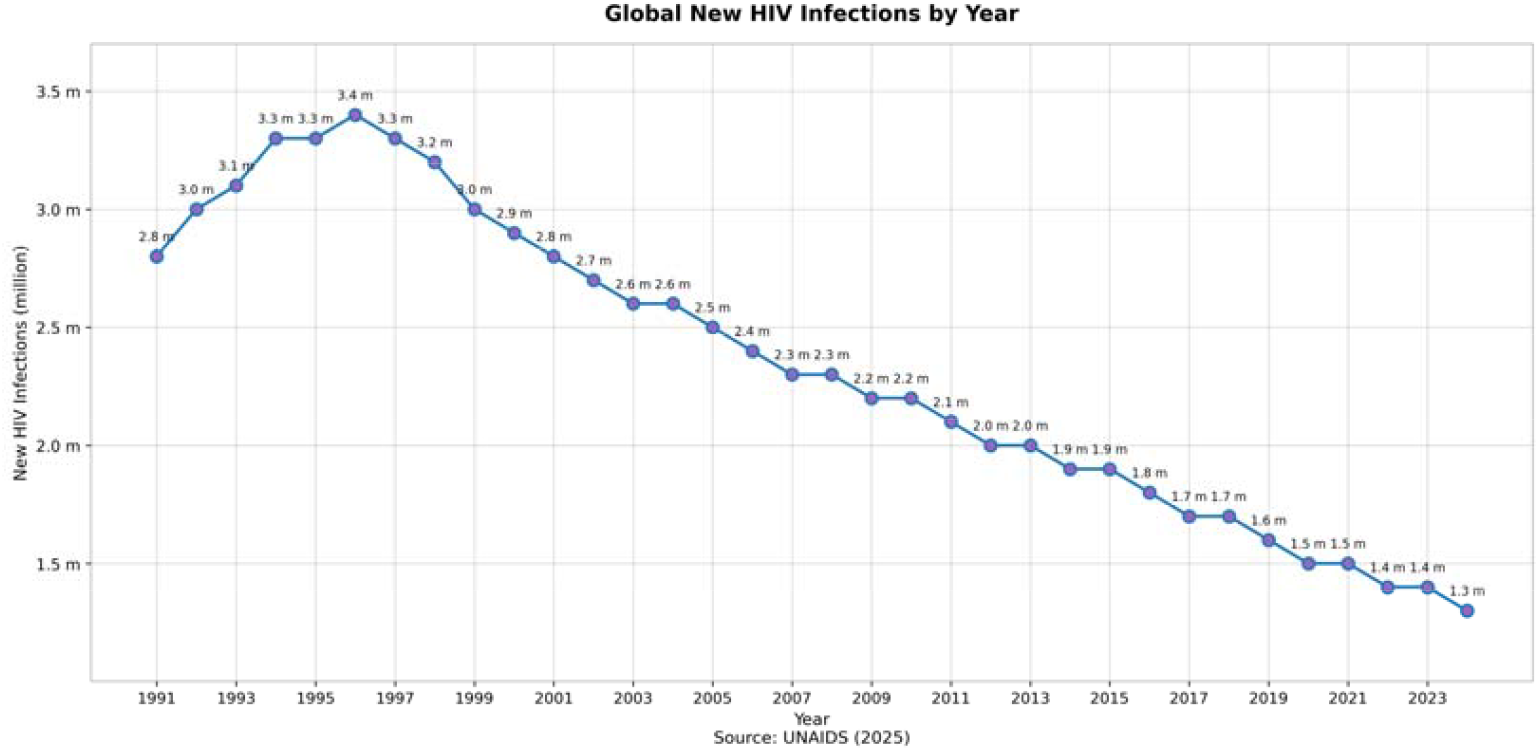
Trends in Global New HIV Infections, 1991–2024.

### 3.2. Global spatial autocorrelation across years (Moran’s I, 2005–2024)

Global Moran’s I is a widely used statistic for assessing global spatial autocorrelation, that is, whether observations exhibit spatial clustering, dispersion, or randomness across geographic units in a given year. It evaluates whether neighboring units tend to show similar values (positive autocorrelation) or dissimilar values (negative autocorrelation) relative to a spatial-randomness null. Statistical significance is commonly judged using the associated z-score and p-value: a significantly positive z-score (e.g., p < 0.05) indicates a non-random clustered pattern, whereas a non-significant p-value suggests the observed spatial pattern does not differ from randomness at the chosen alpha level.

As summarized in Table 1, Moran’s I values were consistently mostly from 2005 to 2024 (range: 0.023–0.302), indicating an overall tendency toward spatial clustering rather than dispersion. Most years showed statistically significant global clustering, for example 2005 (I = 0.180, z = 5.872, p < 0.001) and 2012 (I = 0.302, z = 9.172, p < 0.001). The strongest clustering signal was observed during 2010–2012 (I = 0.251–0.302; z = 5.729–9.172; all p < 0.001). In contrast, the global autocorrelation signal weakened in 2021 (I = 0.023, z = 1.186, p = 0.236; Table X) and became borderline in 2022 (I = 0.045, z = 1.888, p = 0.059), before returning to statistical significance in 2023–2024 (2023: I = 0.068, p = 0.0058; 2024: I = 0.053, p = 0.0349).

**Table 1.**
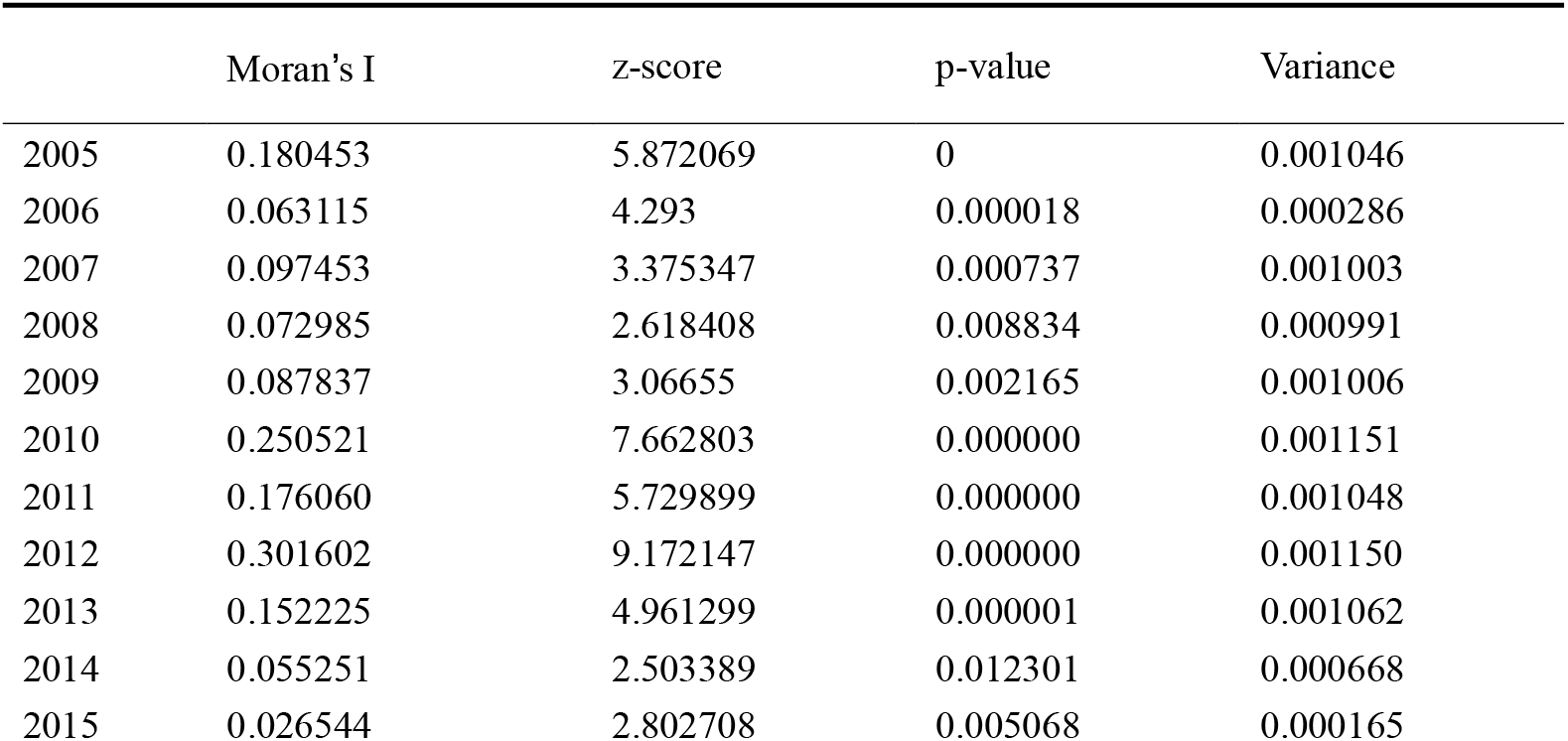
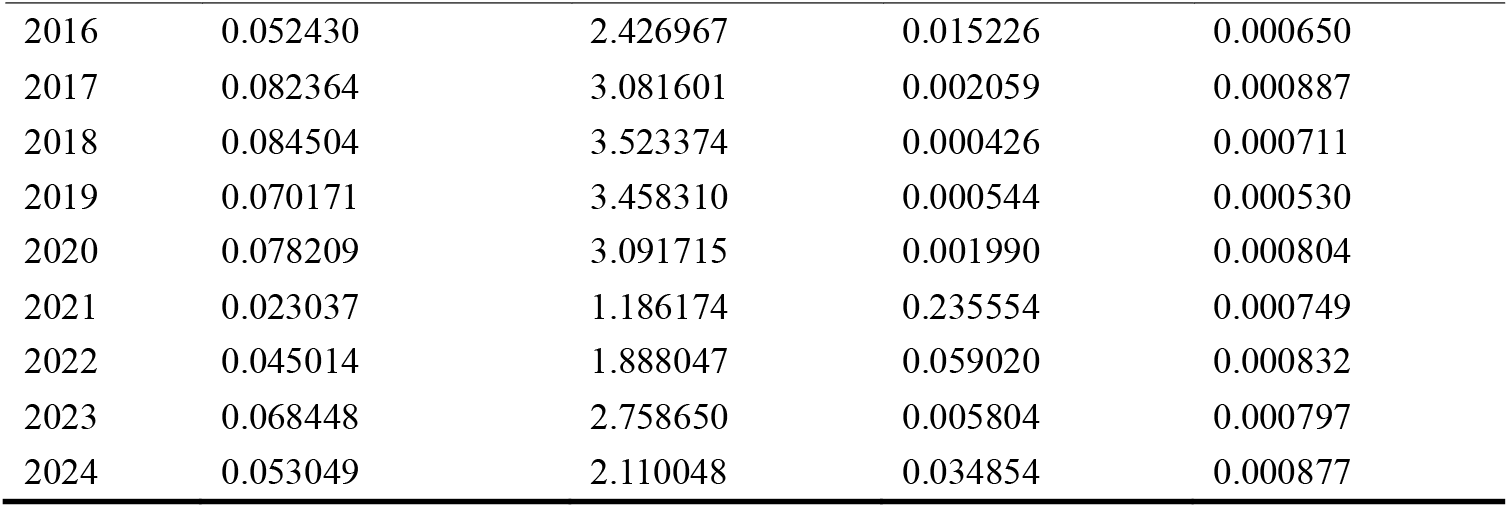
Global Moran’s I Results for HIV-1 Subtype C case Incidence (2005–2024)

|  | Moran's I | z-score | p-value | Variance |
| --- | --- | --- | --- | --- |
| 2005 | 0.180453 | 5.872069 | 0 | 0.001046 |
| 2006 | 0.063115 | 4.293 | 0.000018 | 0.000286 |
| 2007 | 0.097453 | 3.375347 | 0.000737 | 0.001003 |
| 2008 | 0.072985 | 2.618408 | 0.008834 | 0.000991 |
| 2009 | 0.087837 | 3.06655 | 0.002165 | 0.001006 |
| 2010 | 0.250521 | 7.662803 | 0.000000 | 0.001151 |
| 2011 | 0.176060 | 5.729899 | 0.000000 | 0.001048 |
| 2012 | 0.301602 | 9.172147 | 0.000000 | 0.001150 |
| 2013 | 0.152225 | 4.961299 | 0.000001 | 0.001062 |
| 2014 | 0.055251 | 2.503389 | 0.012301 | 0.000668 |
| 2015 | 0.026544 | 2.802708 | 0.005068 | 0.000165 |
| 2016 | 0.052430 | 2.426967 | 0.015226 | 0.000650 |
| 2017 | 0.082364 | 3.081601 | 0.002059 | 0.000887 |
| 2018 | 0.084504 | 3.523374 | 0.000426 | 0.000711 |
| 2019 | 0.070171 | 3.458310 | 0.000544 | 0.000530 |
| 2020 | 0.078209 | 3.091715 | 0.001990 | 0.000804 |
| 2021 | 0.023037 | 1.186174 | 0.235554 | 0.000749 |
| 2022 | 0.045014 | 1.888047 | 0.059020 | 0.000832 |
| 2023 | 0.068448 | 2.758650 | 0.005804 | 0.000797 |
| 2024 | 0.053049 | 2.110048 | 0.034854 | 0.000877 |

Overall, Table 1provides evidence that HIV-1 subtype C sequence counts exhibited statistically significant global spatial clustering in most years, supporting the presence of non-random spatial structure and justifying subsequent local hotspot/coldspot analyses. The non-significant (2021) and marginal (2022) results indicate a temporary attenuation of global clustering at the global scale; however, this does not preclude the existence of localized clusters, which can still be investigated using local indicators of spatial association.

### 3.3. Spatiotemporal descriptive mapping

Quinquennial spatial distribution of subtype C sequence availability. Figure 6 maps the five-year average availability of HIV-1 subtype C sequences standardized by population size (sequences per million population) across four quinquennial periods (2005–2009, 2010–2014, 2015–2019, and 2020–2024). This metric was used to characterize the spatial pattern of sequence availability relative to population size, rather than to represent population-based incidence. Annual sequence counts were standardized by national population estimates and then summarized within each quinquennium to facilitate cross-period visualization of broad spatial shifts.

**Figure 6.**
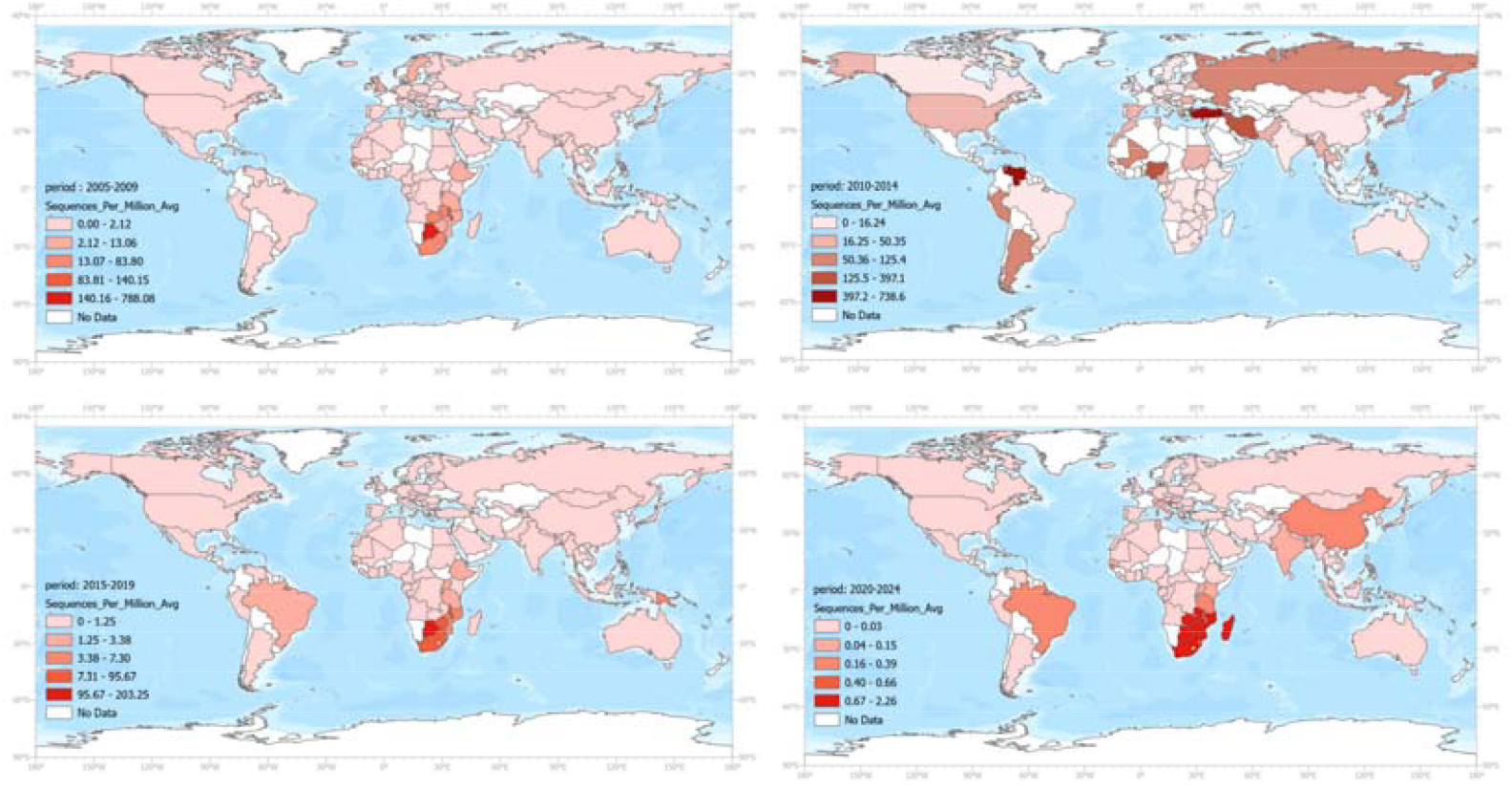
Quinquennial Global Distribution of HIV-1 Subtype C Sequences per Million Population, 2005–2024. (Note: Map lines delineate study areas and do not necessarily depict accepted national boundaries.)

Figure 6 shows that the increase in newly captured sequences in Eastern and Southern Africa remained relatively sustained over the 20-year period compared with other regions, although this dominance appeared slightly weaker during 2010–2014. In addition, during 2010–2014, Greece, Venezuela, Nigeria, and Russia exhibited a modest surge in sequence growth relative to other areas. China and Brazil showed comparatively limited growth during the first 15 years, but experienced a small uptick during 2020–2024 compared with most other countries.

### 3.4. Getis-Ord Gi* results

#### 3.4.1. Classification of Significant Hotspots

In ArcGIS, the Gi_Bin field provides an interpretation of statistical significance based on the Getis-Ord Gi* Z-score and p-value. Each bin corresponds to a specific confidence level for identifying clusters of high (hot spots) or low (cold spots) HIV incidence. The classification used in this study is as follows:(refer to Table 2)

**Table 2.** ArcGIS Gi_Bin Hotspot and Coldspot Significance Classes.

| Gi_Bin | Confidence Level |
| --- | --- |
| +3 | 99% confidence ($p \leq 0.01$) |
| +2 | 95% confidence ($p \leq 0.05$) |
| +1 | 90% confidence ($p \leq 0.10$) |
| 0 | Not significant |
| -1 | 90% confidence ($p \leq 0.10$) |
| -2 | 95% confidence ($p \leq 0.05$) |
| -3 | 99% confidence ($p \leq 0.01$) |

In this study, statistically significant hotspots were defined as Gi_Bin ≥ +2, corresponding to p < 0.05 (95% confidence). Gi_Bin = 3 represented 99% hotspots. Gi_Bin values of −2 and −3 represented significant cold spots

#### 3.4.2. Annual hotspot–coldspot pattern (maps/matrix)

A year-by-year overview of the Getis–Ord Gi* classification was generated for all countries from 2005 to 2024. Figure 7 visualizes the annual Gi_Bin values, allowing inspection of temporal hotspot and cold spot patterns prior to the derivation of the persistence index (the matrix includes only countries that were significant at any point in time).

**Figure 7.**
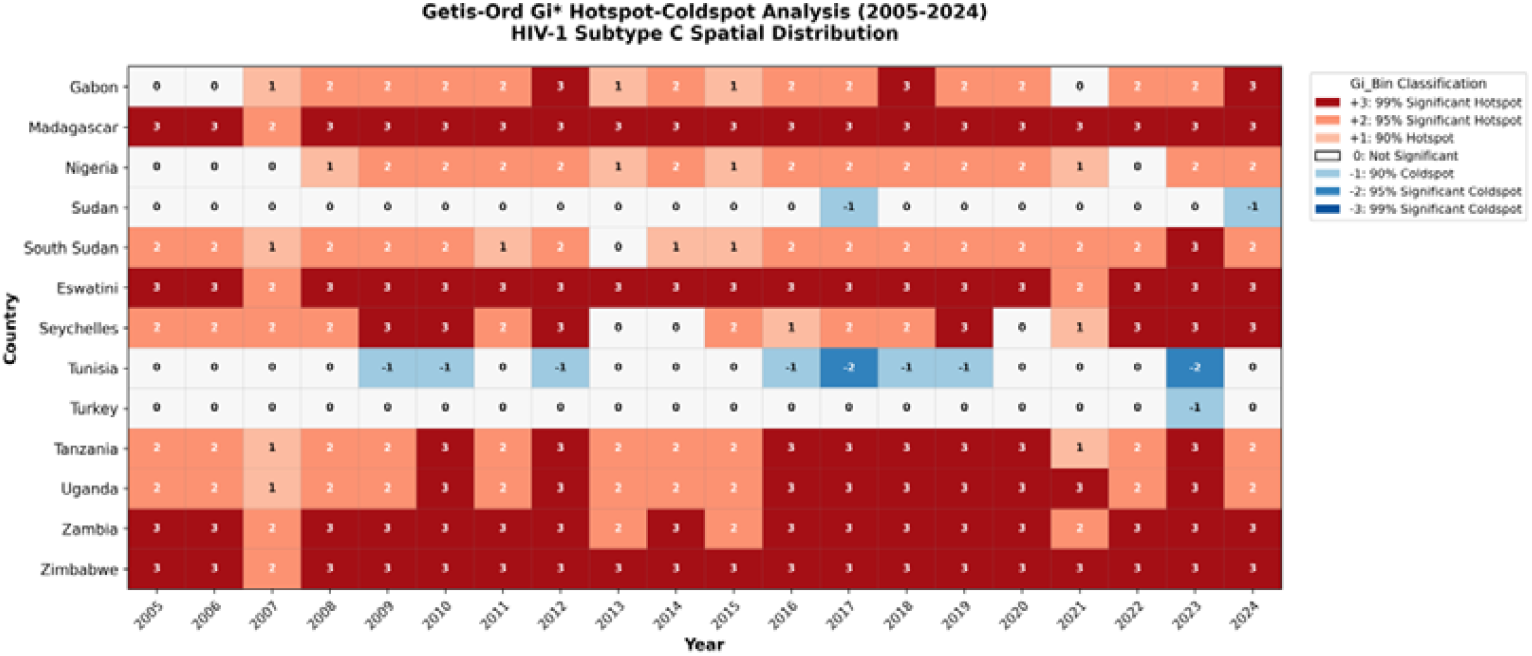
Getis-Ord Gi* Hotspot-coldspot Analysis (2005-2024) HIV-1 Subtype c Spatial Distribution.

The Getis–Ord Gi* hotspot–coldspot matrix revealed clear spatiotemporal variation in the intensity of HIV-1 subtype C activity from 2005 to 2024. Highly significant hotspots (Gi_Bin = +3) persisted across most years in Southern African countries—including Eswatini, Zambia, Zimbabwe, and Madagascar—indicating long-standing, exceptionally elevated transmission intensity. Moderate hotspots (Gi_Bin = +1 to +2) appeared intermittently in East Africa (e.g., Tanzania, Uganda) and Central Africa, reflecting episodic increases in spatial concentration.

Conversely, sporadic coldspot signals (Gi_Bin = −1 to −3) were observed in a small number of countries such as Tunisia and Turkey, suggesting periods of markedly lower-than-expected spatial intensity. Several countries (e.g., Sudan, South Sudan) demonstrated year-to-year fluctuations between neutral and hotspot categories, indicating temporal instability in transmission intensity rather than persistent elevation.

#### 3.4.3. Hotspot Persistence Analysis (2005–2024)

We applied a binary classification to the Getis–Ord Gi* results by recoding the Gi_Bin values such that scores ≥ 2 were assigned a value of 1, while scores < 2 were assigned a value of 0. For each country, the binary values across the 20-year period were summed and divided by 20 to obtain a hotspot stability index. A value closer to 1 indicates a highly stable hotspot, whereas values approaching 0 indicate low stability or inconsistent hotspot patterns (see Table 3 for details).

**Table 3.**
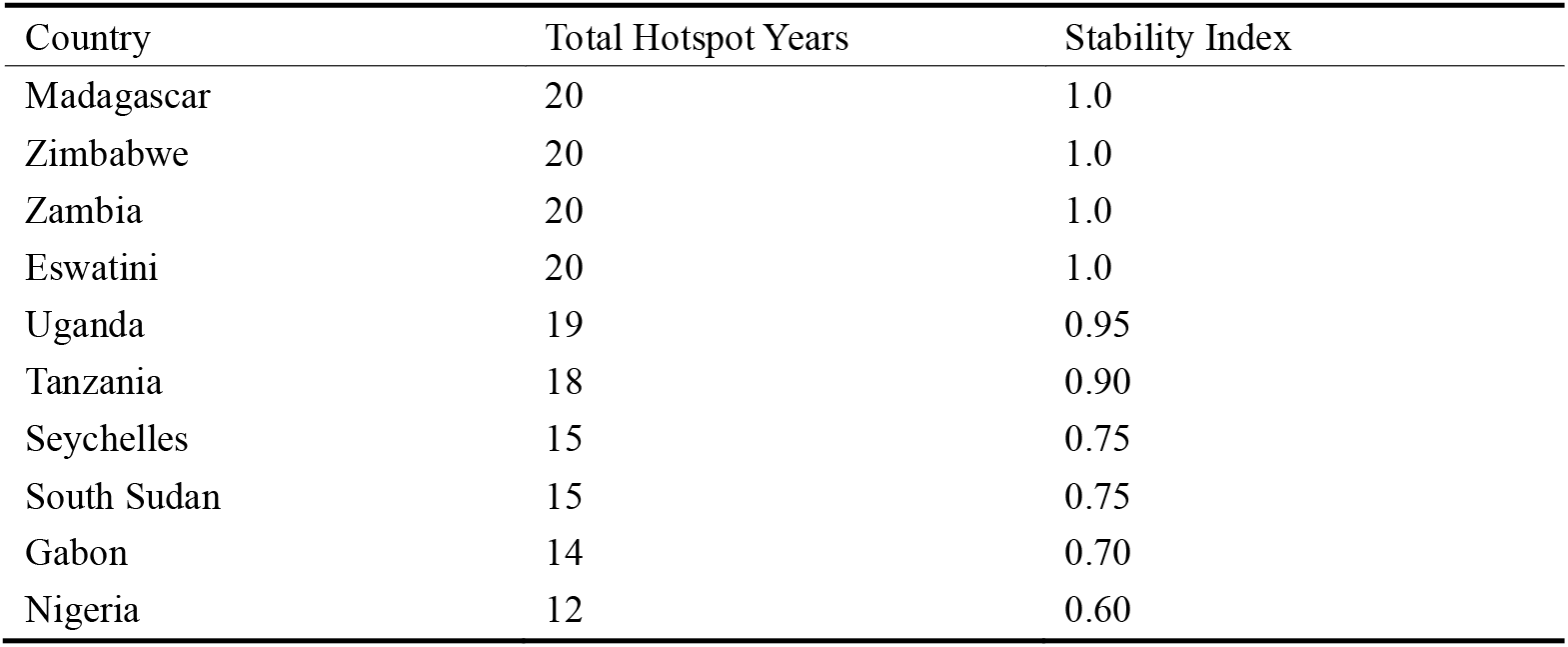
Summary of Hotspot Persistence Based on Binary Gi* Classification (2005–2024)

The resulting stability index was then joined back to the ArcGIS national boundary layer to generate the hotspot persistence map (see Figure 8).

**Figure 8.**
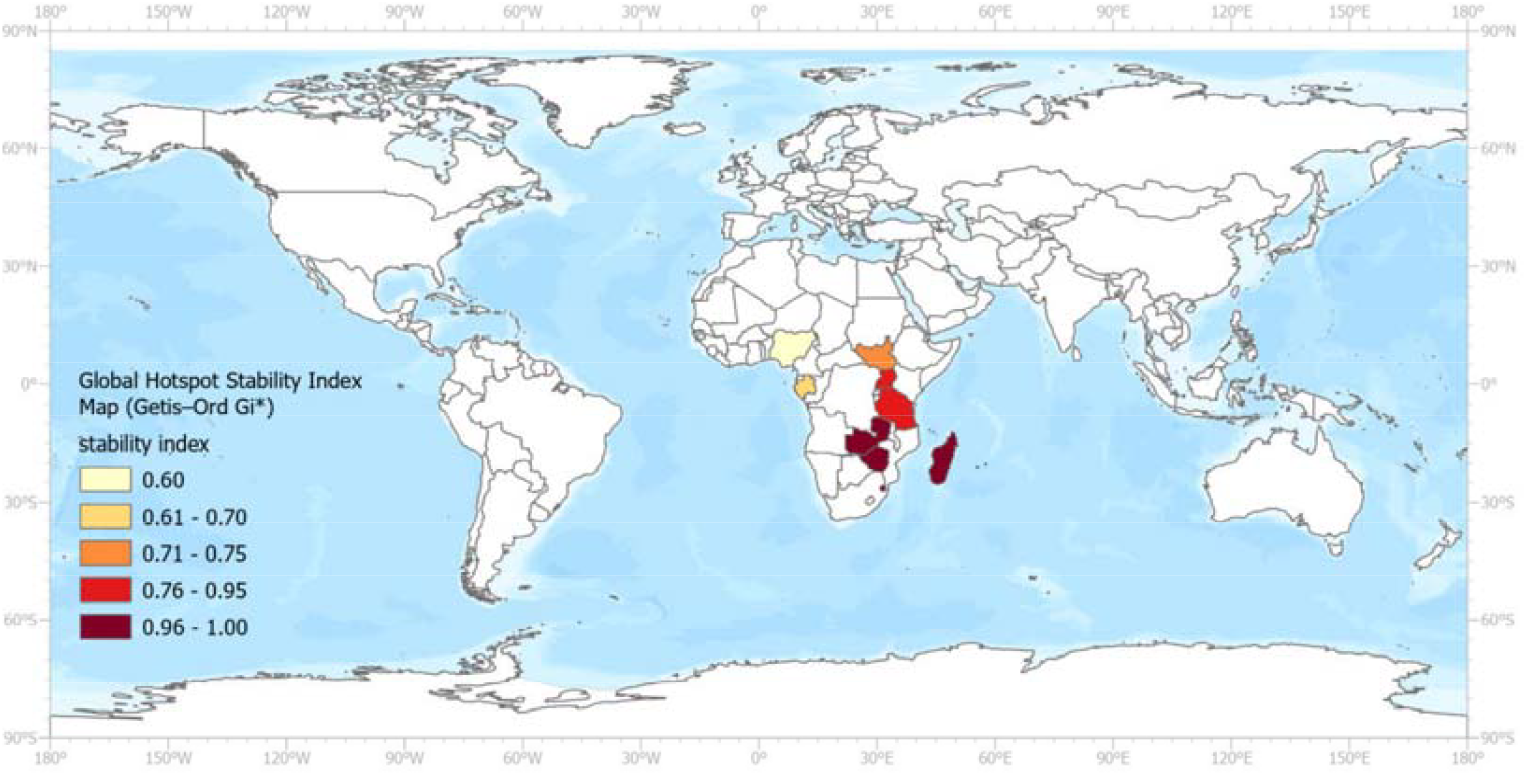
Global Hotspot Stability Index Map (Getis–Ord Gi*) (Note: Map lines delineate study areas and do not necessarily depict accepted national boundaries.)

The Hotspot Stability Index Map (Figure 8) illustrates clear geographic variation in the long-term stability of HIV-1 subtype C hotspots from 2005 to 2024. Southern Africa demonstrates the highest level of persistence, with Madagascar, Zimbabwe, Zambia, and Eswatini exhibiting stability indices of 1.0, indicating that these countries were classified as significant hotspots in every year of the 20-year period. Uganda and Tanzania also show high levels of persistence (0.95 and 0.90, respectively), suggesting consistently elevated hotspot activity with only minor fluctuations.

Intermediate levels of persistence were observed in Seychelles, South Sudan, Gabon, and Nigeria, indicating more intermittent or episodic hotspot activity rather than continuous hotspot persistence.

Overall, the global pattern highlights a concentrated and enduring hotspot zone in Southern Africa, aligning with the dominant epidemiological burden of HIV-1 subtype C in this region. This long-term persistence underscores the entrenched transmission dynamics in these countries and supports the need for sustained, high-intensity prevention strategies.

### 3.5. Bayesian results

#### 3.5.1. Model Fitting and Convergence Diagnostics of Bayesian

The MCMC diagnostic results for the Bayesian BYM2–RW1 model are presented through trace plots that display the iteration-wise trajectories of each Markov chain for the five hyperparameters: α (alpha), (sigma_phi), ρ spatial (rho_spatial), (sigma_gamma), and (sigma_epsilon). Each colored line in Figure 9 represents one of the 64 parallel MCMC chains executed across 1,000 post-tuning iterations.

**Figure 9.**
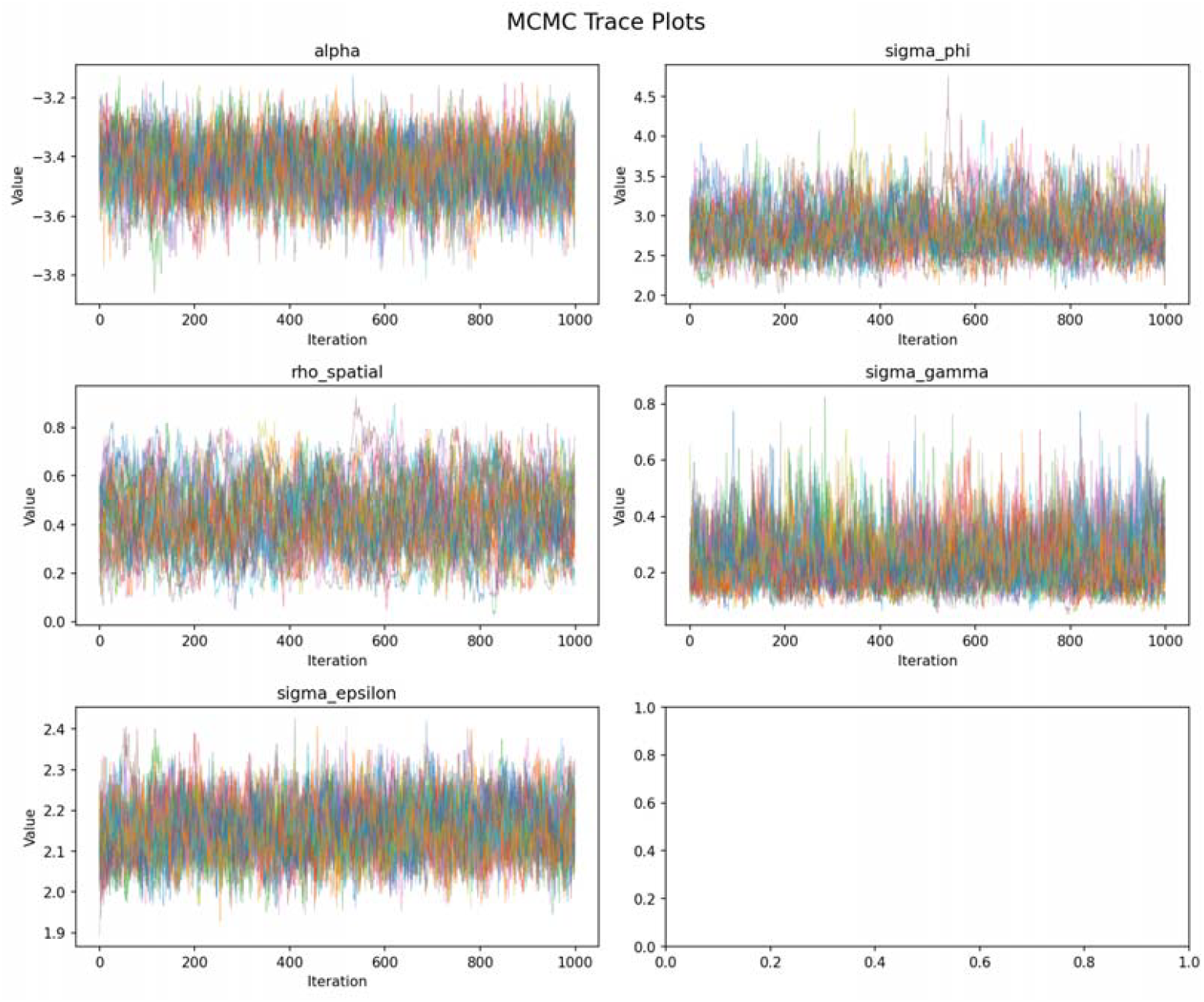
CMC Trace Plots for Hyperparameters of the Bayesian BYM2–RW1 Model.

All parameters exhibit stable horizontal bands without systematic drift, indicating that the chains have reached stationarity and are adequately exploring the posterior distribution. The heavy overlap and interweaving of chains across all parameters further suggest good mixing and the absence of chain-specific modes or trapping behavior.

For the global baseline parameter α, the chains fluctuate consistently between approximately −3.2 and −3.8, showing no long-term drift and indicating a stable estimate of the baseline log-risk. The spatial variance parameter σ_φ_ oscillates within a band between approximately 4.5 and 2.0, reflecting well-identified spatial heterogeneity.

The BYM2 spatial mixing parameter ρ spatial ranges from about 0 to 0.8, with most values concentrated in the 0.2–0.6 range, suggesting that the structured spatial component contributes substantially to overall spatial variation.

The temporal variance parameter σ_γ_ displays stable movement between approximately 0 and 0.8, while the interaction varianceσ_ε_ remains concentrated between about 1.9 and 2.4.

Convergence was further evaluated using the Gelman--Rubin diagnostic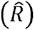. All hyperparameters showed satisfactory convergence, with 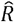 values close to 1 (α ≈ 1.03, σ_φ_≈ 1.03, ρ__spatial__ ≈ 1.05, σ_γ_ ≈ 1.05, and σ_ε_ ≈ 1.02)

The posterior density plots complement the trace plots by illustrating the marginal posterior distributions of the key hyperparameters after convergence. All posterior densities are smooth and unimodal, with no evidence of multimodality or pathological behavior, indicating stable and well-identified parameter estimates. The posterior mean of the intercept α is centered around −3.45, while the spatial standard deviation is concentrated near 2.81, reflecting substantial spatial heterogeneity. The BYM2 spatial mixing parameter (rho_spatial) is centered around 0.43, suggesting a balanced contribution of structured and unstructured spatial effects.

The temporal variance parameter (sigma_gamma) exhibits a mildly right-skewed but unimodal distribution centered around 0.24, consistent with the expected behavior of scale (standard deviation) parameter under weakly informative priors and indicating moderate temporal fluctuations. The space–time interaction variance (sigma_epsilon) is centered near 2.16, highlighting notable local deviations beyond purely spatial or temporal trends. Taken together, the posterior density plots and trace plots demonstrate adequate mixing and convergence of the MCMC chains, providing reliable posterior inference for subsequent estimation of relative risks, hotspot identification, and spatiotemporal cluster analysis (refer to Figure 10).

**Figure 10.**
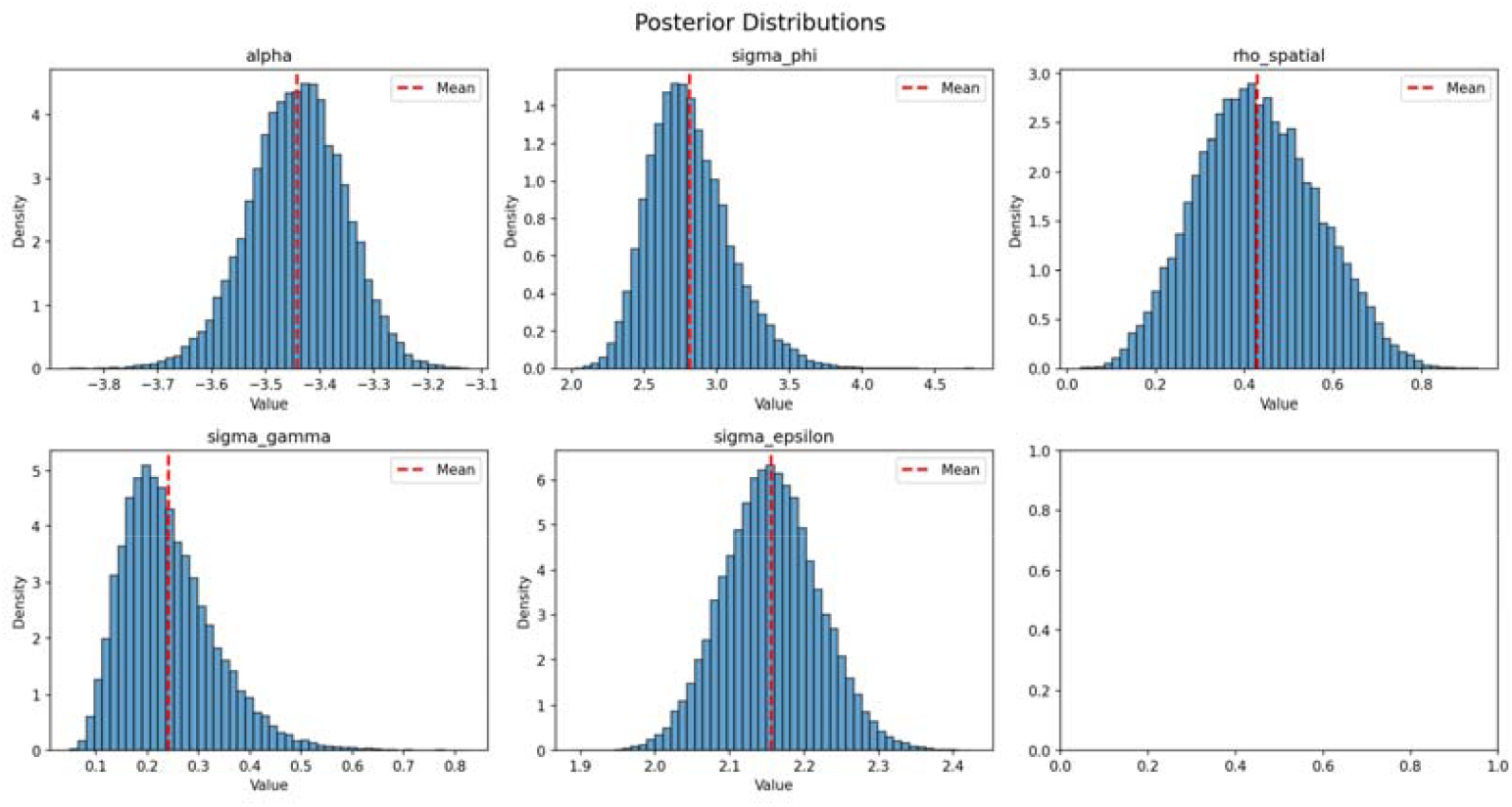
Posterior Distributions of the Hyperparameters in the Bayesian BYM2– RW1 Model.

#### 3.5.2. Classification of Significant Hotspots and Coldpots

To identify high–risk (hotspot) and low–risk (coldspot) areas, we classified each country–year observation based on the model–derived relative risk (RR) and its 95% posterior credible interval.

Specifically, an observation was considered a hotspot if the posterior lower bound of the 95% credible interval exceeded and a coldspot if the posterior upper bound of the 95% credible interval was below.

Using this criterion, all countries that exhibited hotspot or coldspot behaviour in any year were extracted, and their values were organized into a country– by–year matrix. This matrix was subsequently visualized as a heat map, with countries as columns and years as rows, to illustrate the temporal evolution and geographic distribution of coldspots and hotspots (refer to Figure 11).

**Figure 11.**
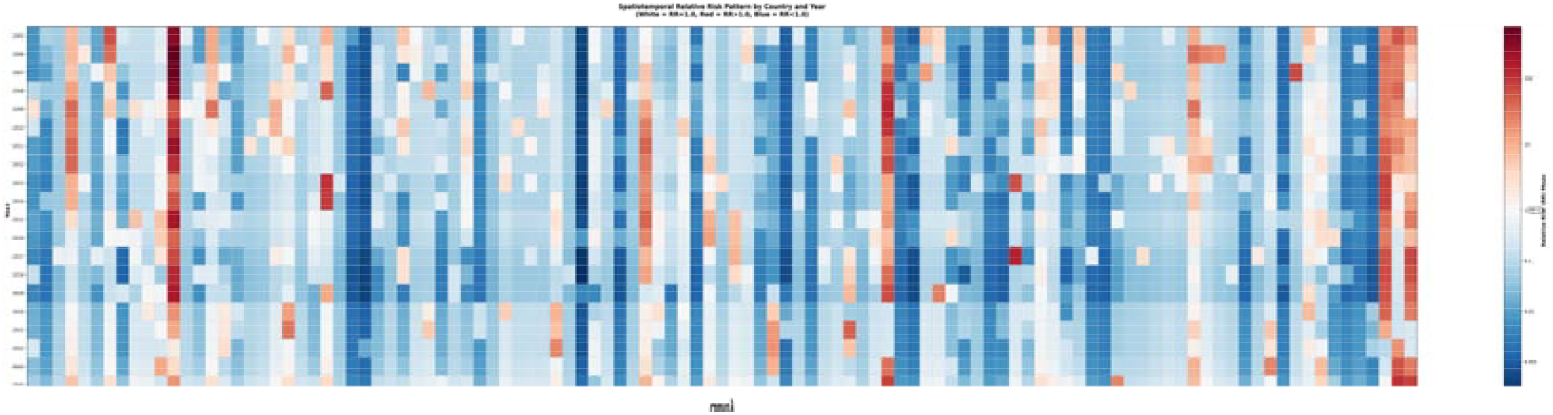
Bayesian Spatiotemporal Relative Risk (RR) Heatmap for HIV-1 Subtype C (2005– 2024)

Analysis of the spatiotemporal RR matrix (Figure 11) revealed several distinct trajectory patterns across countries from 2005 to 2024. A first group of countries, including Australia (AUS), Botswana (BWA), Israel (ISR), Malawi (MWI), and South Africa (ZAF), exhibited a rise–decline–resurgence pattern, characterised by an initial increase in RR values, followed by a marked decline toward or below the baseline, and a subsequent rebound in more recent years.

In contrast, a second group of countries displayed sporadic and fragmented hotspot appearances without sustained temporal continuity. This pattern was observed in countries such as Angola (AGO), Bulgaria (BGR), the Republic of the Congo (COG), Cabo Verde (CPV), Denmark (DNK), Fiji (FJI), Greece (GRC), Kenya (KEN), Liberia (LBR), and several others, reflecting transient elevations in RR that were limited to isolated years.

A third group of countries remained persistent coldspots throughout the study period. Countries including Burundi (BDI), the Dominican Republic (DOM), Egypt (EGY), Spain (ESP), Indonesia (IDN), South Korea (KOR), Mexico (MEX), Malaysia (MYS), Nigeria (NGA), Saudi Arabia (SAU), and Sudan (SDN) consistently exhibited RR values below 1 across nearly all observation years, indicating a sustained lower-than-expected representation of subtype C.

Overall, most countries demonstrated a general tendency toward cooling in recent years, consistent with a broader global decline in subtype C relative risk. Nevertheless, a subset of countries—including Australia (AUS), Botswana (BWA), Switzerland (CHE), Cyprus (CYP), Denmark (DNK), Croatia (HRV), Israel (ISR), Kuwait (KWT), Malawi (MWI), Senegal (SEN), Sweden (SWE), Uganda (UGA), Zambia (ZMB), and Zimbabwe (ZWE)—showed early signs of renewed elevation, suggesting potential localized resurgence dynamics warranting continued surveillance.

#### 3.5.3. Hotspot Persistence Analysis (2005–2024)

In the hotspot persistence analysis, we evaluated the long-term stability of hotspots using the posterior probability that the relative risk exceeds the hotspot threshold,

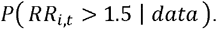

For each country □ that exhibited a hotspot in at least one year, we calculated its 20-year average posterior exceedance probability:

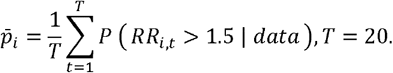

The value 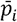 was used to determine whether a hotspot is persistent, moderately persistent, weakly persistent, or non-persistent (see Table 4).

**Table 4.** Classification of Hotspot Persistence Based on the Value of 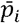.

| Value of $\bar{p}_i$ | Persistence category |
| --- | --- |
| $\bar{p}_i \geq 0.95$ | Extremely persistent hotspot |
| $0.80 \leq \bar{p}_i < 0.95$ | Moderately persistent hotspot |
| $0.50 \leq \bar{p}_i < 0.80$ | Weakly persistent hotspot |
| $\bar{p}_i < 0.50$ | Not persistent |

Based on this index, we constructed a global hotspot persistence summary table (Table 5), which was then imported into ArcGIS to generate the Bayesian hierarchical hotspot persistence map for HIV-1 subtype C. In this map, darker colors denote stronger and more persistent hotspots, whereas lighter colors indicate unstable or weakly persistent hotspots (refer to Figure 12).

**Table 5.** Posterior mean exceedance probability 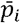 for all hotspot countries (2005–2024).

| Country | Average $\bar{p}_i$ | Persistence_category |
| --- | --- | --- |
| BWA | 0.944185938 | Moderately persistent hotspot |
| ZAF | 0.8630234375000001 | Moderately persistent hotspot |
| MWI | 0.850232813 | Moderately persistent hotspot |
| ZWE | 0.838671875 | Moderately persistent hotspot |
| ISR | 0.722665625 | Weakly persistent hotspot |
| SWE | 0.704371875 | Weakly persistent hotspot |
| ZMB | 0.6396468750000001 | Weakly persistent hotspot |
| AUS | 0.61971875 | Weakly persistent hotspot |
| DNK | 0.4062249999999999 | Not persistent |
| CYP | 0.391229688 | Not persistent |
| TZA | 0.386757813 | Not persistent |
| PRT | 0.372601563 | Not persistent |
| CHE | 0.339490625 | Not persistent |
| BEL | 0.324998438 | Not persistent |
| MOZ | 0.314101563 | Not persistent |
| ETH | 0.301907813 | Not persistent |
| KWT | 0.2942015624999999 | Not persistent |
| UGA | 0.272973438 | Not persistent |
| ROU | 0.253715625 | Not persistent |
| GBR | 0.24570625 | Not persistent |
| HRV | 0.231490625 | Not persistent |
| LKA | 0.2211875 | Not persistent |
| CUB | 0.214395313 | Not persistent |
| BRA | 0.2125625 | Not persistent |
| MDG | 0.200996875 | Not persistent |
| SWZ | 0.173015625 | Not persistent |
| NOR | 0.1560875 | Not persistent |
| CHN | 0.15 | Not persistent |
| TUN | 0.140946875 | Not persistent |
| NLD | 0.124654688 | Not persistent |
| PNG | 0.116629688 | Not persistent |
| BGR | 0.11644375 | Not persistent |
| FJI | 0.115771875 | Not persistent |
| RWA | 0.111495313 | Not persistent |
| NPL | 0.1009421874999999 | Not persistent |
| SEN | 0.100379688 | Not persistent |
| LUX | 0.093165625 | Not persistent |
| ITA | 0.06908125 | Not persistent |
| MNG | 0.068225 | Not persistent |
| KEN | 0.063460938 | Not persistent |
| SYC | 0.0567078124999999 | Not persistent |
| TWN | 0.055725 | Not persistent |
| IRL | 0.055601563 | Not persistent |
| SGP | 0.052148438 | Not persistent |
| AGO | 0.0502531249999999 | Not persistent |
| USA | 0.050004688 | Not persistent |
| LBR | 0.0497437499999999 | Not persistent |
| SLE | 0.047864063 | Not persistent |
| IND | 0 | Not persistent |

**Figure 12.**
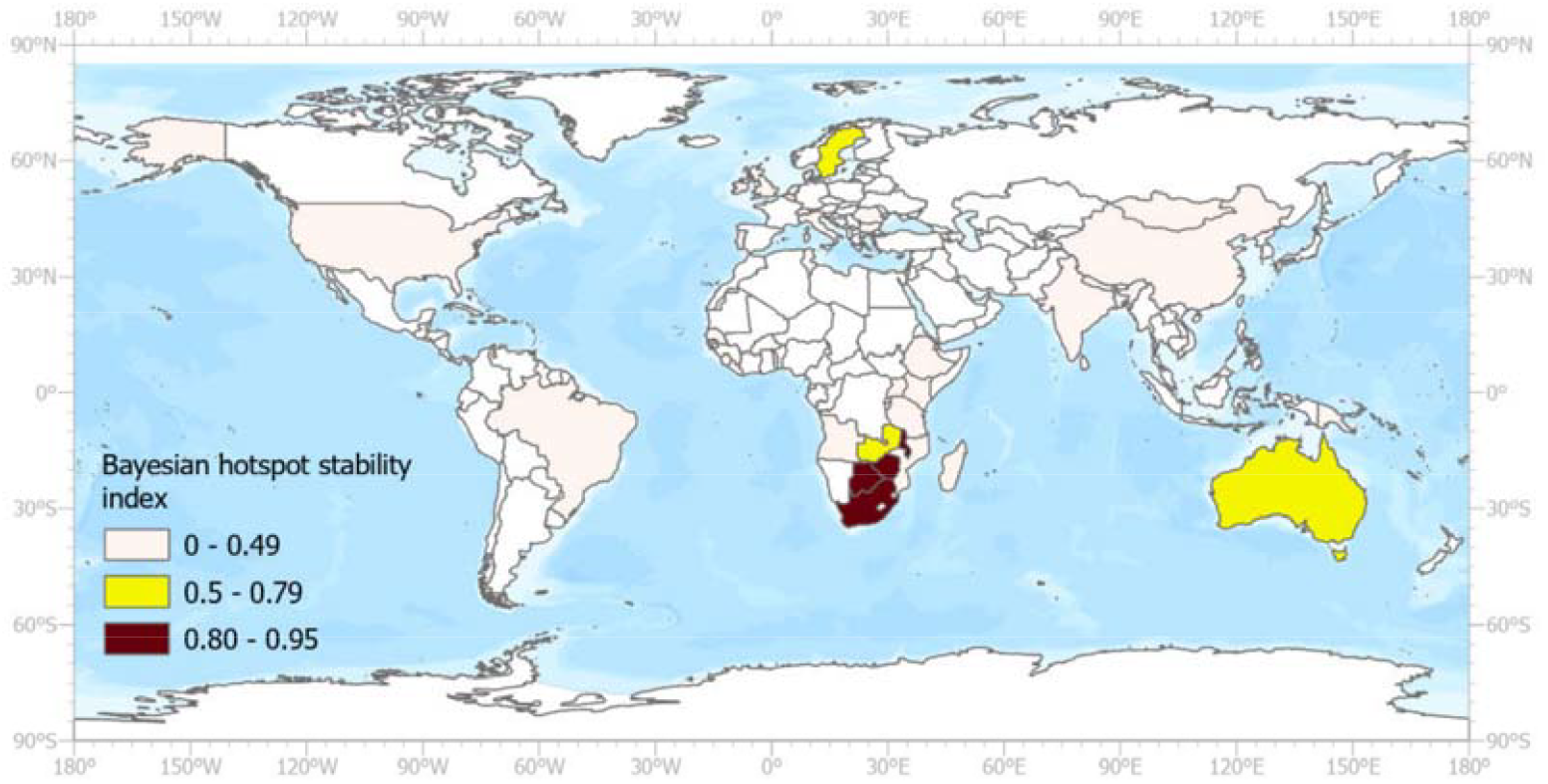
Bayesian Hotspot Persistence Map Based on Posterior Probability 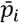 (2005-2024) (Note: Map lines delineate study areas and do not necessarily depict accepted national boundaries.)

### 3.6. SaTScan results

Spatiotemporal scan statistics identified 25 statistically significant clusters of HIV-1 subtype C (P < 0.05) during 2005-2024. The most likely cluster (Cluster 1) was located in Southern Africa, spanning six countries (Eswatini, Botswana, Zimbabwe, South Africa, Mozambique, and Malawi) from 2010 to 2019, with a spatial radius of 1,504 km and a relative risk of 105.65 (74,906 observed vs. 1,314.83 expected cases, P < 0.001). Cluster 2 overlapped geographically but occurred earlier (2005-2008), encompassing seven countries including Zambia (RR = 78.27, 31,999 cases, P < 0.001). Together, these clusters demonstrated sustained high-intensity transmission over 15 years.

Additional clusters were identified in Brazil (Cluster 3: 2015-2017, RR = 4.55), Sudan/Ethiopia (Clusters 4, 6: 2005-2010 and 2017-2018, RR = 3.52 and 3.98), and Northern Europe (Cluster 5: 2006-2014, RR = 2.78, spanning Norway, Denmark, Sweden, and the United Kingdom). Several smaller clusters exhibited notably high relative risks despite limited geographic scope, including Papua New Guinea (Cluster 7: 2017, RR = 30.25, 312 cases; Cluster 10: 2013, RR = 12.95, 121 cases) and Fiji (Cluster 22: 2008, RR = 15.70, 16 cases). The remaining clusters (9-25) were predominantly single-year or short-duration events reflecting more transient transmission dynamics. Three additional clusters were detected but did not reach statistical significance (Clusters 26-28, P > 0.05), suggesting potential areas of interest for future surveillance.

Cluster shapefiles generated by SaTScan were imported into ArcGIS Pro for visualization and spatial analysis (refer toFigure 13).

**Figure 13.**
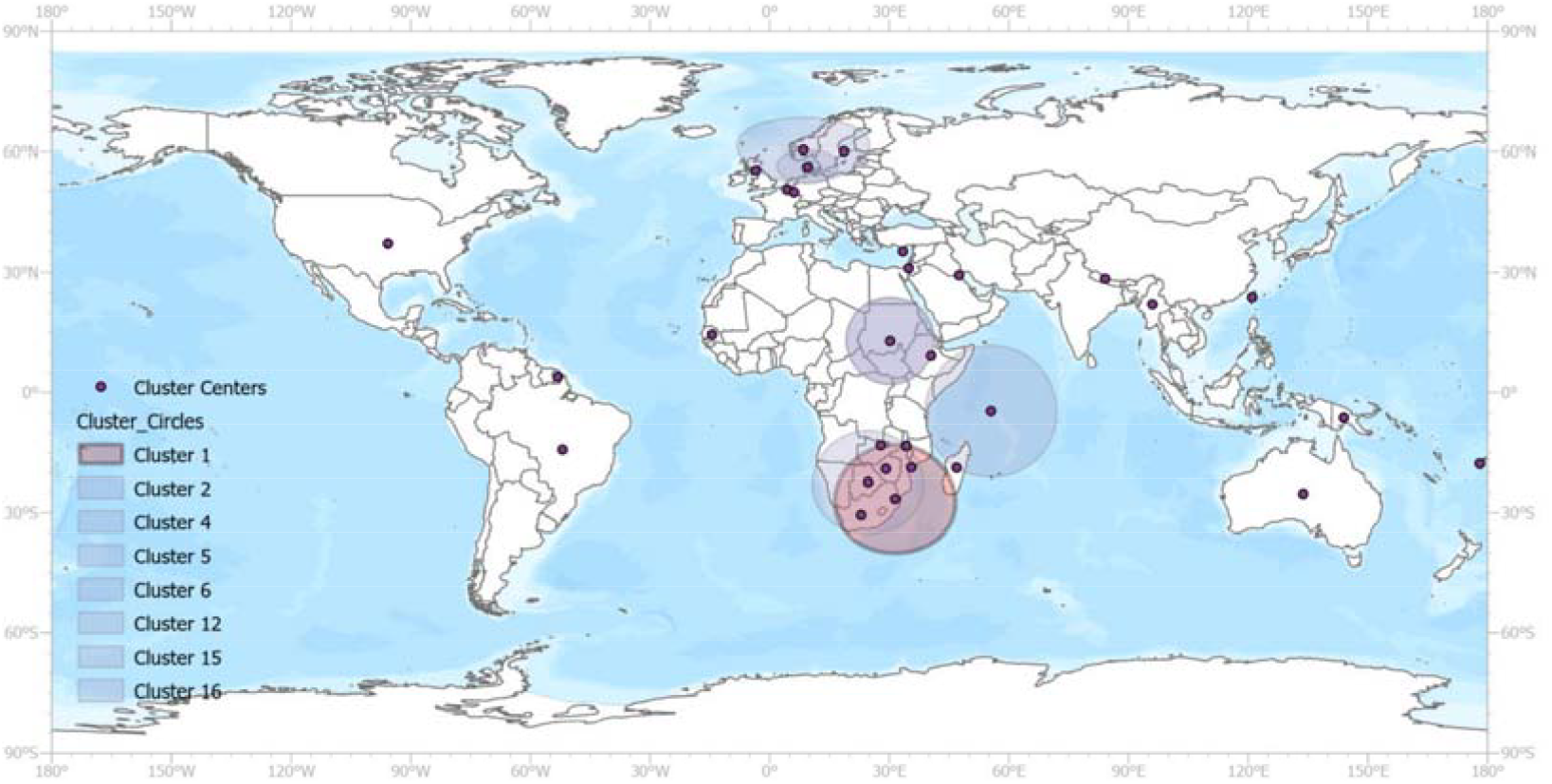
Significant Spatiotemporal Clusters of HIV-1 Subtype C Identified by SaTScan (2005–2024) (Note: Map lines delineate study areas and do not necessarily depict accepted national boundaries.)

#### 3.6.1. Classification of Significant Hotspots

A country was classified as a hotspot if it appeared in any statistically significant spatiotemporal cluster (P < 0.05) identified by SaTScan. The matrix displays countries (vertical axis) across calendar years (horizontal axis), with each colored cell indicating cluster membership in that year.

White cells represent years without significant clustering(refer Figure 14 and Table A 1).

**Figure 14.**
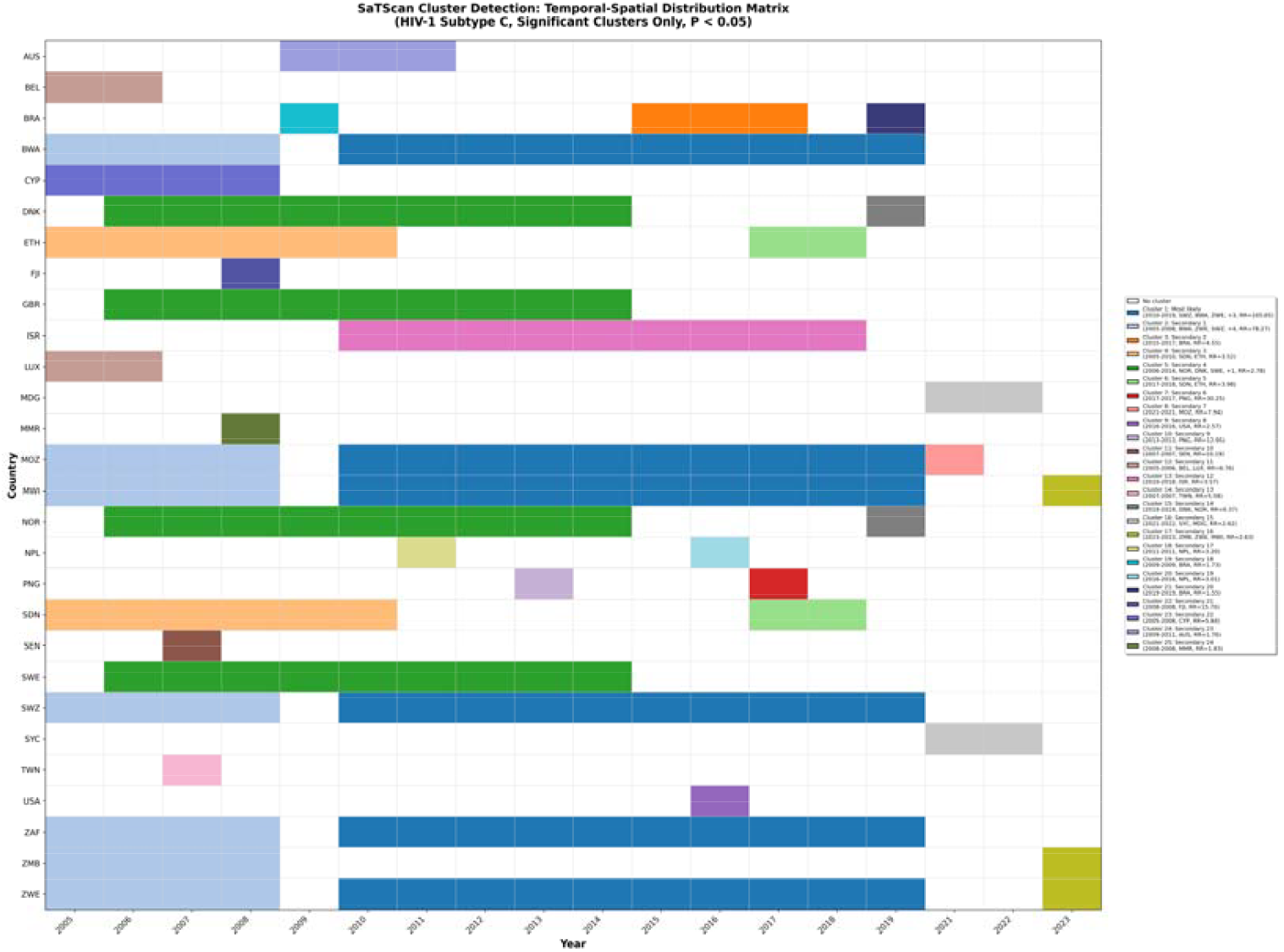
SaTscan Spatiotemporal Hotspot Timeline Matrix (2005-2024): Country-Level Cluster Participation.

The visualization reveals marked heterogeneity in hotspot timing and duration. Southern African countries (Eswatini [SWZ], Botswana [BWA], Zimbabwe [ZWE], Mozambique [MOZ], Malawi [MWI], South Africa [ZAF]) exhibited sustained hotspot periods spanning 14-15 years (2005-2019), driven by two overlapping major clusters (Cluster 1: 2010-2019, RR=105.65; Cluster 2: 2005-2008, RR=78.27), reflecting nearly 15 years of high-intensity transmission. Northern European countries (Norway [NOR], Denmark [DNK]) demonstrated moderate stability with 10-year hotspot involvement. By contrast, countries such as Brazil [BRA], Papua New Guinea [PNG], Senegal [SEN], Taiwan [TWN], and the United States [USA] showed brief, isolated hotspot episodes of 1-5 years. The matrix also illustrates temporal overlap—particularly in Southern Africa where consecutive clusters involved the same countries—highlighting the complex spatiotemporal layering of the epidemic over two decades.

#### 3.6.2. Hotspot Persistence Analysis (2005–2024)

To assess the temporal persistence of hotspot involvement, we calculated the stability percentage for each country, defined as the proportion of years (within 2005–2024) during which that country appeared in a statistically significant spatiotemporal cluster. Based on this stability percentage, countries were classified into three levels: High Stability (≥80%), Moderate Stability (50–79%), and Low Stability (<50%). This classification enables the differentiation between countries experiencing long-term, consistent hotspot activity and those with sporadic or short-lived clustering.

Based on the stability analysis, countries involved in spatiotemporal clusters exhibited marked heterogeneity. Southern African countries — Eswatini (SWZ), Botswana (BWA), Zimbabwe (ZWE), Mozambique (MOZ), Malawi (MWI), and South Africa (ZAF)—demonstrated Moderate Stability (70-75%), with hotspot involvement spanning 14-15 years of the 20-year study period. This persistent pattern was driven primarily by two large overlapping clusters (Cluster 1: 2010-2019; Cluster 2: 2005-2008) (refer to Table 6 and Table A 1).

**Table 6.** Country-Level Hotspot Stability Based on SaTScan Spatiotemporal Cluster Membership (2005– 2024)

| Countr<br>y | Hotspot_Ye<br>ars | Total_Yea<br>rs | Stability_Percent<br>age | Stability_Le<br>vel | Num_Clust<br>ers | Cluster_I<br>Ds |
| --- | --- | --- | --- | --- | --- | --- |
| MWI | 15 | 20 | 75 | Moderate | 3 | 1,2,17 |
| ZWE | 15 | 20 | 75 | Moderate | 3 | 1,2,17 |
| MOZ | 15 | 20 | 75 | Moderate | 3 | 1,2,8 |
| BWA | 14 | 20 | 70 | Moderate | 2 | 1,2 |
| SWZ | 14 | 20 | 70 | Moderate | 2 | 1,2 |
| ZAF | 14 | 20 | 70 | Moderate | 2 | 1,2 |
| NOR | 10 | 20 | 50 | Moderate | 2 | 5,15 |
| DNK | 10 | 20 | 50 | Moderate | 2 | 5,15 |
| GBR | 9 | 20 | 45 | Low | 1 | 5 |
| SWE | 9 | 20 | 45 | Low | 1 | 5 |
| ISR | 9 | 20 | 45 | Low | 1 | 13 |
| ETH | 8 | 20 | 40 | Low | 2 | 4,6 |
| SDN | 8 | 20 | 40 | Low | 2 | 4,6 |
| ZMB | 5 | 20 | 25 | Low | 2 | 2,17 |
| BRA | 5 | 20 | 25 | Low | 3 | 3,19,21 |
| CYP | 4 | 20 | 20 | Low | 1 | 23 |
| AUS | 3 | 20 | 15 | Low | 1 | 24 |
| MDG | 2 | 20 | 10 | Low | 1 | 16 |
| LUX | 2 | 20 | 10 | Low | 1 | 12 |
| BEL | 2 | 20 | 10 | Low | 1 | 12 |
| NPL | 2 | 20 | 10 | Low | 2 | 18,20 |
| SYC | 2 | 20 | 10 | Low | 1 | 16 |
| PNG | 2 | 20 | 10 | Low | 2 | 7,10 |
| FJI | 1 | 20 | 5 | Low | 1 | 22 |
| SEN | 1 | 20 | 5 | Low | 1 | 11 |
| MMR | 1 | 20 | 5 | Low | 1 | 25 |
| TWN | 1 | 20 | 5 | Low | 1 | 14 |
| USA | 1 | 20 | 5 | Low | 1 | 9 |

Northern European countries—Norway (NOR) and Denmark (DNK)—also showed Moderate Stability (50%), participating in clusters for 10 years, primarily within Cluster 5 (2006-2014) and Cluster 15 (2019). Their hotspot patterns were more episodic but indicated sustained transmission during discrete periods.

In contrast, the majority of countries (n=20) demonstrated Low Stability (<50%), with hotspot participation ranging from 1-9 years. Countries such as Ethiopia (ETH), Sudan (SDN), and Brazil (BRA) exhibited intermediate involvement (5-8 years, 25-40%), while others including Papua New Guinea (PNG), Taiwan (TWN), United States (USA), and Senegal (SEN) showed minimal participation (1-2 years, 5-10%). These short-duration hotspots reflect more localized or transient increases in HIV-1 subtype C transmission rather than continuous epidemic intensity.

Notably, no country achieved High Stability (≥80%), underscoring the temporal variability of cluster membership even in persistently affected regions. The stability classification highlights a core region of moderate-stability countries in Southern Africa.

Using SaTScan output, we calculated for each country the percentage of the 20-year period during which it participated in any spatiotemporal cluster (with overlapping clusters in the same year counted once). These values were mapped in ArcGIS to generate the HIV-1 subtype C hotspot stability map (Figure 15).

**Figure 15.**
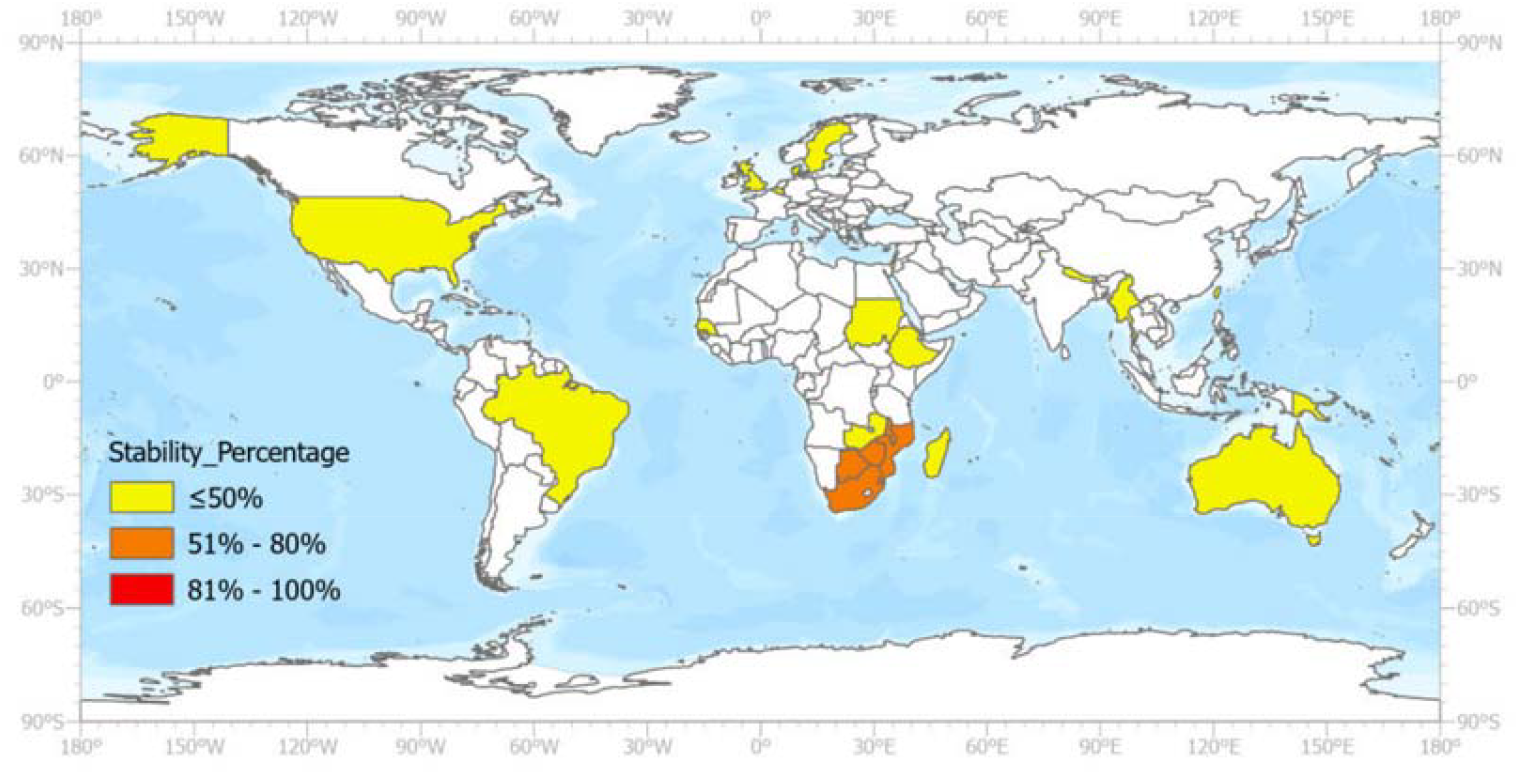
Spatial distribution of hotspot stability for HIV-1 subtype C Case based on SaTScan-identified clusters, 2005–2024. (Note: Map lines delineate study areas and do not necessarily depict accepted national boundaries.)

### 3.7. Comparative analysis

#### 3.7.1. Comparative Analysis of Hotspot Identification Across Three Models

Based on the comparative analysis across three spatial detection methodologies, substantial heterogeneity emerged in hotspot identification patterns. GiBin detected the largest unique set (108 country-year combinations), reflecting its high sensitivity to localized, year-specific clustering through the Getis-Ord Gi* statistic. Bayesian models detected 74 unique combinations, capturing moderate-risk areas through hierarchical spatial-temporal smoothing. SaTScan identified 42 unique hotspots, primarily coherent regional clusters in Northern Europe, Mozambique (MOZ), and Sudan (SDN)(refer to Table A 2**Error! Reference source not found**.).

Cross-model concordance analysis revealed 24 country-year combinations detected by all three methods— representing the most methodologically robust hotspots. These stable detections concentrated overwhelmingly in Southern Africa: Zimbabwe (ZWE, 14 years: 2005-2007, 2010-2019, 2023), Zambia (ZMB, 5 years: 2005-2008, 2023), Eswatini (SWZ, 3 years: 2006, 2011-2012), and Madagascar (MDG, 2 years: 2021-2022). Zimbabwe’s sustained detection across consecutive periods (2005-2007, 2010-2019) indicates structurally entrenched high-risk transmission patterns for HIV-1 subtype C.

Pairwise model overlap varied substantially. Bayesian and SaTScan shared the most extensive concordance (105 country-year pairs), predominantly in Botswana (BWA), Malawi (MWI), South Africa (ZAF), and Mozambique (MOZ) across extended periods, reflecting shared sensitivity to persistent, spatially coherent patterns. Bayesian-GiBin shared 28 combinations, while GiBin-SaTScan overlap was minimal (13 pairs). The geographic concentration of cross-model stable hotspots in Southern Africa aligns with established epidemiological evidence of subtype C dominance in this region, reinforcing the external validity of our comparative analytical framework.

To facilitate visual interpretation, we constructed a spatiotemporal matrix with years on the vertical axis and countries (ISO A3 codes) on the horizontal axis, assigning distinct colors to each detection model (Figure 16). This visualization framework enables systematic comparison of hotspot evolution patterns and methodological concordance across the 2005-2024 study period.

**Figure 16.**
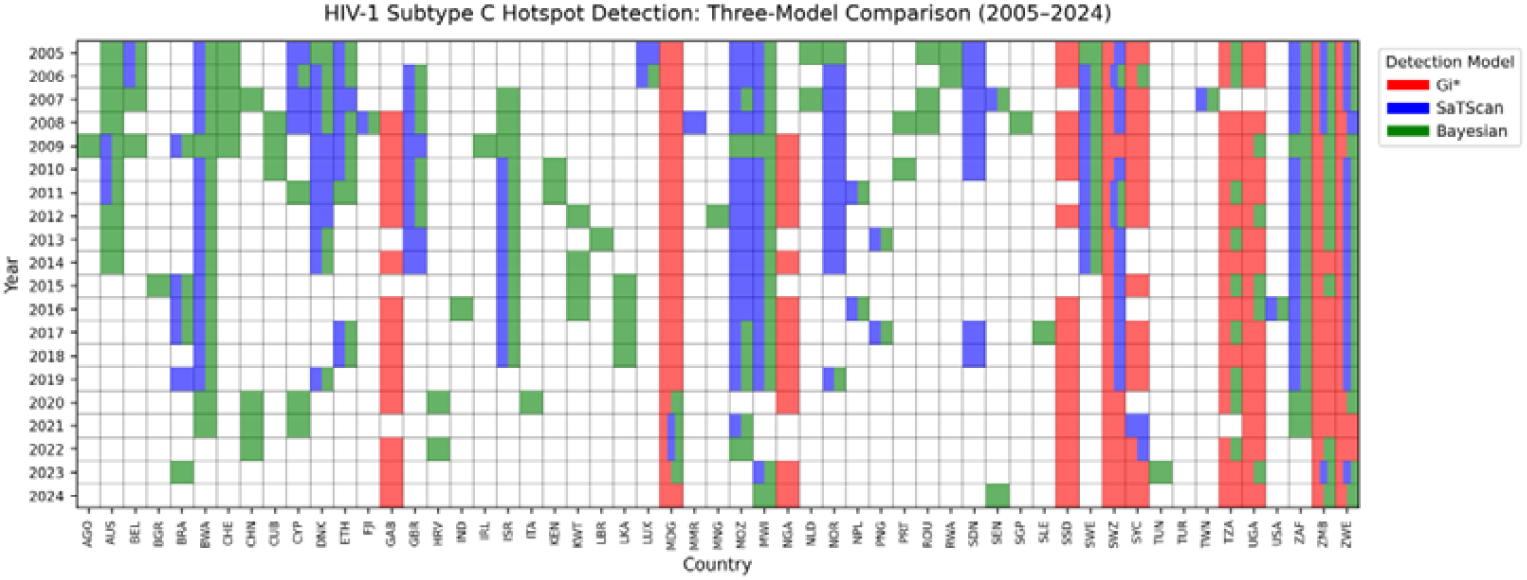
Country–Year Matrix Showing Hotspot Detections by Gi, SaTScan, and Bayesian Models (2005– 2024)

#### 3.7.1. Hotspot Stability Categorization Summary

The hotspot stability—defined as the average number of years in which a country was identified as a hotspot over the 20-year study period—was classified into three levels based on predefined thresholds: high stability (≥0.6), medium stability (0.5–0.6), and low stability (<0.5). Using this scheme, we compared stability values generated by the three analytical models (Gi*, Bayesian, and SaTScan) and organized countries into mutually exclusive categories, including: (i) hotspots consistently identified as highly stable across all models, (ii) hotspots recognised as highly stable by two models, (iii) hotspots considered highly stable by only one model, and (iv) locations that fall within the medium or low stability ranges. This classification framework allows the shared and model-specific components of hotspot stability to be clearly identified (refer to Table 7).

**Table 7.**
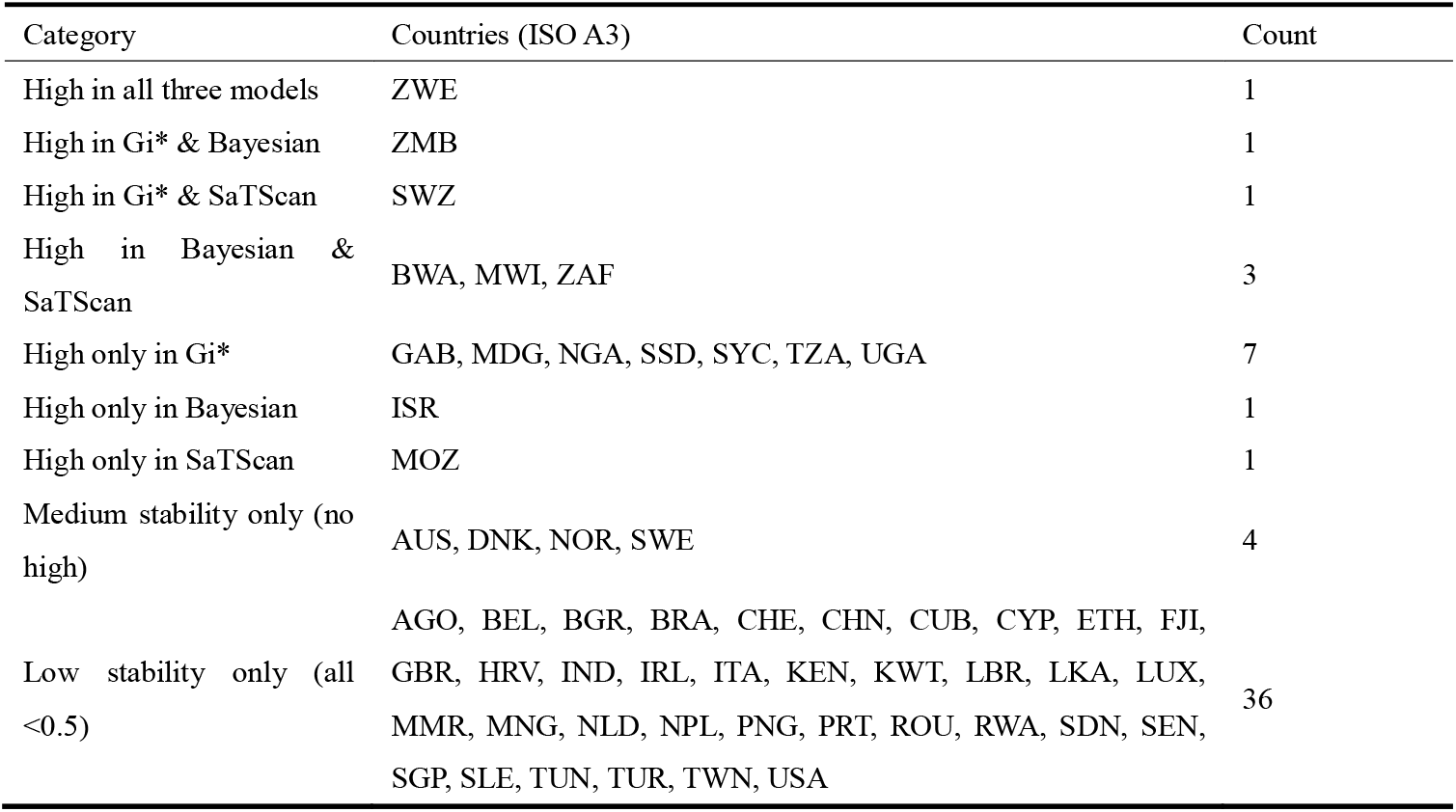
Categorization of Countries by Hotspot Stability Across Three Detection Models.

Zimbabwe (ZWE) was the only country classified as a high-stability hotspot across all three detection models, indicating methodologically robust and sustained HIV-1 subtype C transmission over the 20-year study period.

Several countries achieved high-stability classification in two models. Zambia (ZMB) was identified by both Gi* and Bayesian methods, while Eswatini (SWZ) was detected by Gi* and SaTScan. Botswana (BWA), Malawi (MWI), and South Africa (ZAF) were jointly identified by Bayesian and SaTScan models.

Model-specific high-stability hotspots revealed distinct methodological sensitivities. Seven countries—Gabon (GAB), Madagascar (MDG), Nigeria (NGA), South Sudan (SSD), Seychelles (SYC), Tanzania (TZA), and Uganda (UGA)—were identified exclusively by Gi*. Israel (ISR) achieved high stability only in the Bayesian model, while Mozambique (MOZ) was identified exclusively by SaTScan. Four countries—Australia (AUS), Denmark (DNK), Norway (NOR), and Sweden (SWE)—showed medium stability only, without achieving high stability in any model. The remaining 36 countries exhibited low stability across all models.

For visualization, countries ever identified as hotspots were displayed on the horizontal axis with detection models on the vertical axis (Figure 17). Cell colors represented model-specific stability: red (Gi*), green (Bayesian), and blue (SaTScan), with intensity reflecting the proportion of years classified as a hotspot.

**Figure 17.**
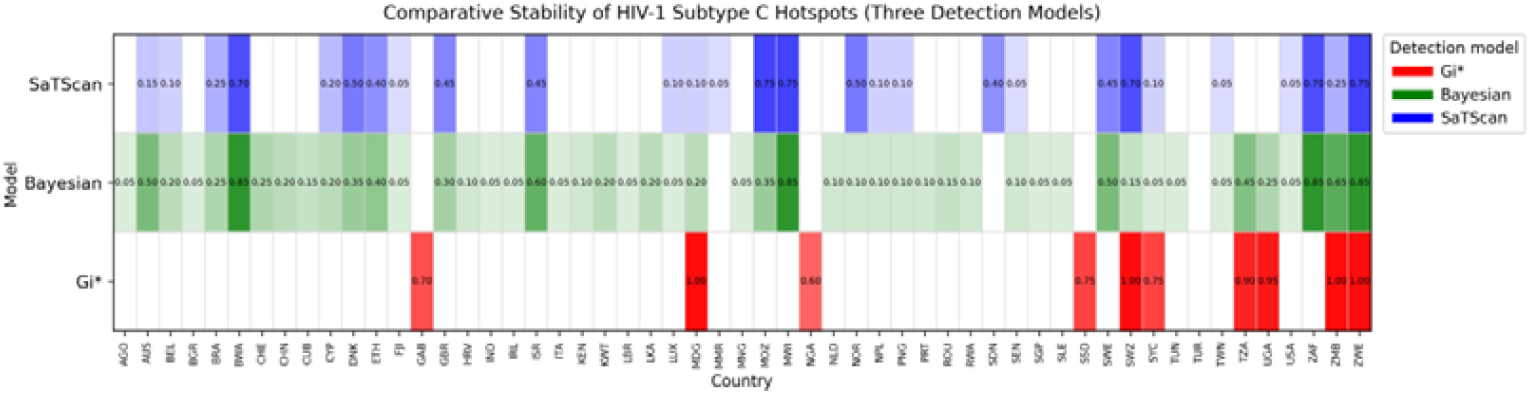
Matrix Visualization of Hotspot Stability Across Three Detection Models.

## 4. Discussion

Research on non-B HIV subtypes is a relatively novel field. Historically, in Western countries, HIV-1 subtype B has been the predominant circulating subtype. Possibly due to economic and research-priority considerations, most studies have therefore focused on subtype B. However, non-B subtypes constitute the dominant share of HIV infections globally, and subtype C is the most prevalent among the non-B lineages. Previous geographical studies of subtype C have largely relied on descriptive report or have applied GIS-based clustering tests within limited regions. In contrast, GIS-based spatial clustering analyses of HIV-1 subtype C at the global scale remain largely absent.

In the context of accelerating globalization, international population mobility is becoming increasingly close and frequent, and the restrictive effect of national borders on human movement is gradually weakening. Accordingly, global-scale GIS clustering research on HIV-1 subtype C is becoming progressively more important. Moreover, HIV subtypes differ substantially in replicative capacity, preferentially affected populations, immune escape potential, and propensity for drug resistance. Therefore, subtype-specific geographical analyses can provide meaningful guidance for global HIV molecular surveillance and source tracing, drug-resistance monitoring, resource allocation, and vaccine development.

### 4.1. Main finding

Analysis of 20-year sequence data from the LANL HIV Database revealed a clear transition in the global distribution of HIV-1 subtype C from widespread geographic dispersion toward concentrated spatial persistence. Documented hotspots evolved from scattered multi-continental distribution in the early period (2005–2014) to progressively localized concentration in Southern Africa during the later period (2015–2024). This spatial consolidation pattern, characterized by a shift from diffuse surface-level transmission to focused point-source persistence, was consistently detected across all three analytical methods, indicating robust evidence for geographic convergence of subtype C transmission activity.

The three spatial analytical approaches demonstrated distinct but complementary detection characteristics. Getis-Ord Gi* exhibited high sensitivity to short-term spatial anomalies and strong dependence on neighboring country data for local spatial autocorrelation calculation, resulting in hotspot identification that was highly concentrated within contiguous Southern African regions. In contrast, Bayesian hierarchical modeling and SaTScan, both incorporating spatiotemporal smoothing mechanisms, detected additional hotspots beyond the Southern African core, including episodic elevated transmission in China, India, and Australia during specific years. The absence of temporal smoothing in Gi* annual analyses produced greater year-to-year variability in hotspot classification, contributing to substantial divergence from the other two methods. This methodological sensitivity gradient reflects fundamental trade-offs between detecting localized spatial clustering (Gi*) versus identifying temporally stable transmission patterns (Bayesian/SaTScan).

Zimbabwe (ZWE) emerged as the only country achieving high-stability hotspot classification across all three independent analytical methods, demonstrating exceptional cross-method robustness as a core transmission hub. This convergent identification, despite each method’s distinct statistical frameworks, assumptions, and sensitivity profiles, provides strong evidence for Zimbabwe’s persistent role in sustaining regional subtype C transmission networks. Four additional Southern African countries achieved high-stability classification in two methods: Zambia (ZMB) by Gi* and Bayesian, Eswatini (SWZ) by Gi* and SaTScan, and Botswana (BWA), Malawi (MWI), and South Africa (ZAF) by Bayesian and SaTScan. These five countries form a geographically contiguous “Southern African transmission corridor” centered on Zimbabwe. The multi-method convergence observed exclusively in this region underscores its epidemiological significance as the sustained epicenter of documented HIV-1 subtype C activity.

The country-year relative risk (RR) matrix derived from Bayesian hierarchical modeling revealed a declining trend in global HIV-1 subtype C hotspot intensity over the past decade (2015–2024). Although Southern African countries maintained elevated RR values throughout the study period, no country sustained RR(low)> 1.0 across all 20 years, and most non-core regions exhibited marked RR declines after 2015. This temporal pattern suggests a global cooling of documented subtype C transmission intensity, potentially reflecting genuine epidemiological trends (such as enhanced prevention and treatment programs), shifts in genomic surveillance capacity, or changing research priorities in sequence deposition. Notably, even within persistent hotspot countries, RR values demonstrated temporal fluctuation rather than sustained elevation, indicating dynamic epidemic phases rather than static endemic equilibrium.

By comparing the combined hotspot matrix derived from the three models(Figure 16) with the hotspot matrices generated by the Bayesian(Figure 11) and SaTScan(Figure 14)approaches, it becomes evident that although HIV-1 subtype C still remains the predominant subtype globally, its peak period has passed. However, this pattern is not as apparent in the Getis–Ord Gi* country– year hotspot matrix(Figure 7). This discrepancy may stem from the fact that SaTScan filters out random fluctuations through spatial scanning windows and Monte Carlo significance testing, while the Bayesian model incorporates spatial–temporal smoothing that prevents brief or minor variations from being classified as hotspots. In contrast, the Getis–Ord Gi* statistic relies mainly on local spatial values and lacks comparable filtering mechanisms, causing its well-known limitations to become more pronounced.

Geographic visualization of high-stability hotspots revealed a predominantly African distribution, with the majority concentrated in Eastern and Southern Africa. Notably, all countries achieving high-stability classification in two or more models (Zimbabwe, Zambia, Eswatini, Botswana, Malawi, South Africa) were exclusively Southern African, forming a spatially contiguous transmission corridor (Figure 18).

**Figure 18.**
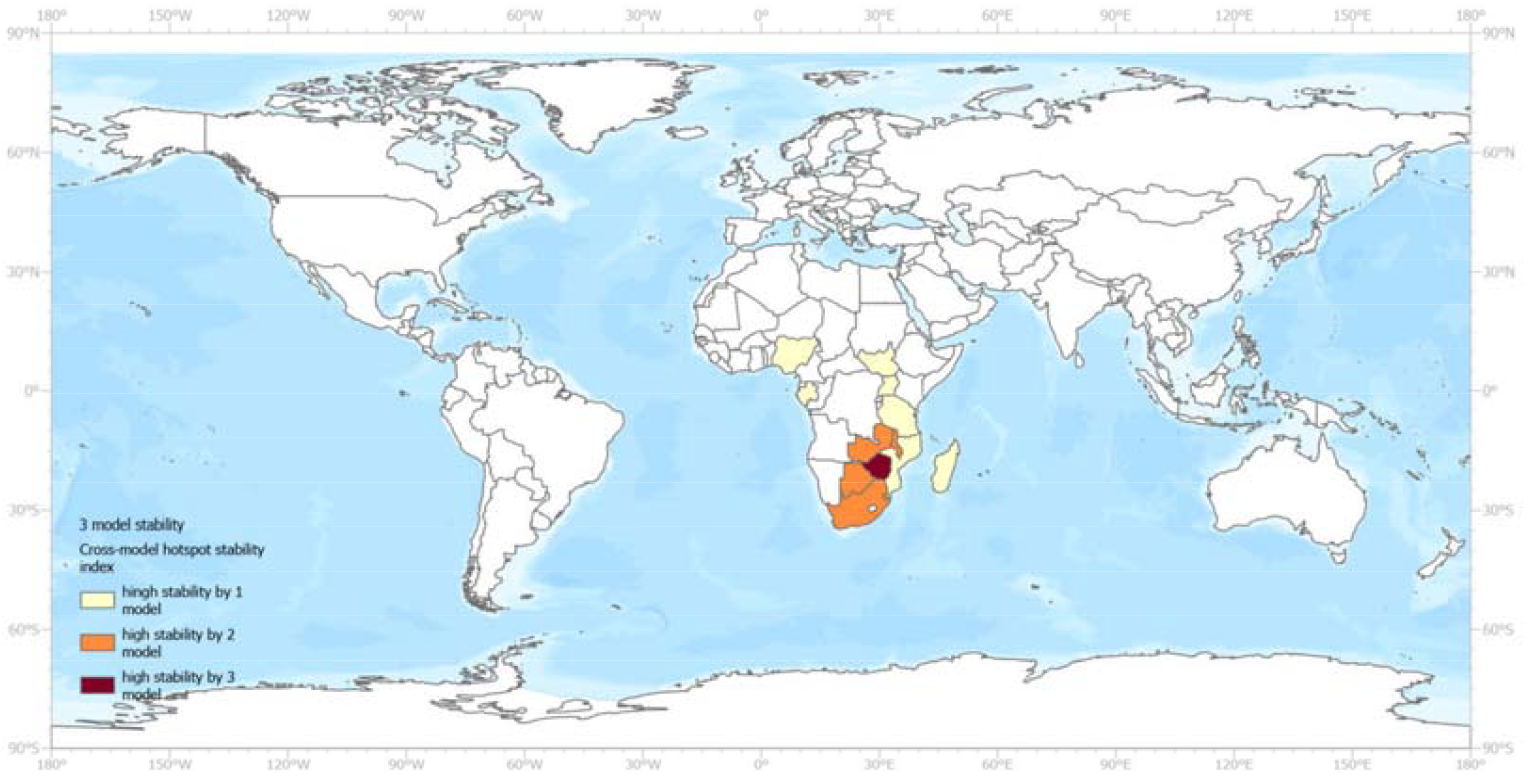
Geographic clustering of high-stability HIV-1 subtype C hotspots by the level of model agreement (1–3 models) (Note: Map lines delineate study areas and do not necessarily depict accepted national boundaries.)

### 4.2. Comparison with Previous Studies

Comparison with previously published geographic distributions of HIV-1 subtype C revealed varying levels of concordance across the three analytical methods. The Bayesian hierarchical model exhibited the broadest geographic coverage, identifying hotspots in Southern Africa, Eastern Africa, India and neighboring countries, and southwestern China, consistent with established epidemiological patterns (Giovanetti et al., 2020; Oka et al., 2019; Chen et al., 2019).

In comparison, Getis-Ord Gi* identified hotspots exclusively within Eastern and Southern Africa, while SaTScan detected clusters in India and neighboring countries but did not classify China as a significant hotspot. These discrepancies likely reflect methodological differences in handling population-weighted incidence rates: both China and India have exceptionally large populations, which may dilute per-capita case counts and reduce sensitivity in methods that rely on absolute spatial clustering (Gi*) or fixed-radius cluster detection (SaTScan). The Bayesian approach, which incorporates spatial random effects and temporal smoothing, appears less susceptible to population size effects and therefore captured a broader geographic distribution more consistent with known epidemiological patterns.

Based on the comparative results from the three models, we summarized the strengths and limitations of each approach Table 8.

**Table 8.** Comparison of Spatial Detection Methods.

| Method | Strengths | Limitations |
| --- | --- | --- |
| Getis-Ord $G_i^*$ | High sensitivity to localized spatial clustering; effective for detecting year-specific anomalies; no temporal assumptions required | High year-to-year variability; cannot assess long-term persistence; susceptible to sampling artifacts; dependent on neighboring countries |
| SaTScan | Conservative with low false-positive rate; identifies persistent spatiotemporal clusters; robust statistical testing; filters random fluctuations | Lower sensitivity to diffuse hotspots; may miss gradual spatial diffusion; cylinder-based detection constrains cluster shapes; population size effects |
| Bayesian<br>BYM2-RW1 | Comprehensive spatiotemporal assessment; borrows information across space-time; stabilizes estimates in data-sparse areas; less affected by population size; predictive capability | May respond to small fluctuations; smoothing may dampen localized signals; cannot overcome surveillance gaps; computationally intensive |

### 4.3. Public Health Implications

Our findings provide actionable guidance for optimizing HIV-1 genomic surveillance and resource allocation. Zimbabwe’s unique status as the only country achieving high-stability hotspot classification across all three independent methods justifies its prioritization as a sentinel surveillance site for early detection of drug resistance mutations, novel recombinant forms, and regional transmission network dynamics. The identified Southern African transmission corridor (Zimbabwe, Zambia, Eswatini, Botswana, Malawi, South Africa) underscores the need for coordinated regional surveillance rather than isolated national programs, given cross-border population mobility and interconnected transmission networks. Beyond this high-priority corridor, a tiered approach is warranted: moderate-intensity monitoring in regions with episodic hotspot patterns (India, China) and strategic sampling elsewhere to detect potential re-emergence events. This stratified allocation optimizes limited surveillance resources while maintaining global situational awareness.

### 4.4. Limitations

This study’s primary limitation stems from reliance on HIV-1 sequence data from the LANL public database, which reflects genomic surveillance capacity rather than comprehensive epidemiological burden. Sequencing density varies substantially across countries: nations with robust laboratory infrastructure (e.g., South Africa, Botswana, high-income settings) contribute disproportionately more sequences relative to case burden, while many high-prevalence, resource-limited regions remain underrepresented. This surveillance heterogeneity means that identified hotspots represent documented transmission clustering in available sequences rather than definitive epidemiological prevalence patterns across all infections. To partially address this limitation, we employed a Bayesian hierarchical spatiotemporal model incorporating BYM2 (Besag–York–Mollié) spatial random effects (Riebler et al., 2016). This specification partitions spatial variation into structured (spatially autocorrelated via Gaussian Markov Random Field) and unstructured (independent) components, enabling information sharing across neighboring regions through precision-weighted smoothing. Combined with year-specific standardization (ensuring within each year, ΣExpected = ΣObserved) and population offset terms in the Poisson log-linear framework, this approach stabilizes risk estimates in data-sparse areas and reduces artifacts from extreme population heterogeneity. However, spatial smoothing represents an inherent methodological trade-off: while attenuating false signals driven by sampling noise, it may also dampen detection of genuinely localized transmission hotspots that differ sharply from spatially proximate regions. Moreover, no statistical model can reconstruct information absent from the data—Bayesian methods redistribute uncertainty more evenly but cannot overcome fundamental surveillance gaps. Therefore, findings should be interpreted as characterizing the spatial structure of available genomic surveillance rather than providing definitive epidemiological risk maps. Future research integrating sequence data with case-based surveillance, behavioral surveys, and systematic representative sampling would strengthen inference about true transmission patterns.

HIV-1 subtype C sequences were retrieved from the Los Alamos National Laboratory (LANL) HIV Sequence Database. A single-database strategy was adopted for three reasons. First, WHO and UNAIDS do not provide a systematic country–year dataset on HIV-1 subtype distributions. Second, LANL’s sequence records are sourced from NCBI GenBank, and GenBank exchanges nucleotide data with other major repositories through the International Nucleotide Sequence Database Collaboration (INSDC) and related inter-database agreements; therefore, aggregating sequences across multiple repositories or platforms would be methodologically redundant and would increase the risk of duplicate entries. Third, although LANL ultimately relies on GenBank as the primary submission archive, LANL provides an analysis-ready resource with expert curation and standardized subtype annotation, applying additional annotation and quality-control procedures (Kuiken et al., 2003). Collectively, these considerations support the use of LANL as the sole data source for this study.

### 4.5. Conclusions

This study demonstrates that HIV-1 subtype C has transitioned from widespread distribution toward concentrated persistence in Southern Africa, with Zimbabwe uniquely identified as a high-stability hotspot across three independent methods. Systematic comparison reveals complementary capabilities: Gi* for localized clustering, SaTScan for discrete outbreaks, and Bayesian modeling for comprehensive assessment with highest concordance to established endemic regions. These findings inform strategic surveillance prioritization, with the Southern African corridor warranting enhanced monitoring. The integrative framework is transferable to other pathogens, and future integration with epidemiological data will strengthen transmission network inference and evidence-based control strategies.

## Data Availability

The data used in this study were obtained from publicly available, aggregated, and de-identified sources. The original data sources are described in the manuscript. The processed data and related analysis materials are currently stored in a Google Drive folder and are available at the link provided below.

https://docs.google.com/document/d/1bre95LlVGX1gwmC0Iwe3_GEB-lwuRxss/edit

## Finding

This research did not receive any specific grant from funding agencies in the public, commercial, or not-for-profit sectors

## Declaration of generative AI and AI-assisted technologies in the manuscript preparation process

Statement: During the preparation of this work, the authors used ChatGPT and Claude to assist with language editing and improving readability, to support content organization, and to help draft code for data processing after the Getis-Ord Gi* and Bayesian analyses were completed. After using these tools, the authors reviewed, verified, and edited all outputs as needed and take full responsibility for the content of the published article.

## Data availability statement

The HIV-1 subtype C sequence data analysed in this study were obtained from the Los Alamos National Laboratory (LANL) HIV Sequence Database and are subject to the database’s data access policies. Derived datasets generated during the current study, including country–year aggregated counts, hotspot classification results, stability metrics, and all analysis scripts, are publicly available at: [<u>link</u>]. World Bank population data and UNAIDS HIV incidence estimates used for rate standardisation are publicly accessible from their respective official websites.

## Appendices A

Table A1 provides a summary of the raw SaTScan spatiotemporal cluster outputs, which directly correspond to and complement the patterns shown in Figure 14.

**Table A1.**
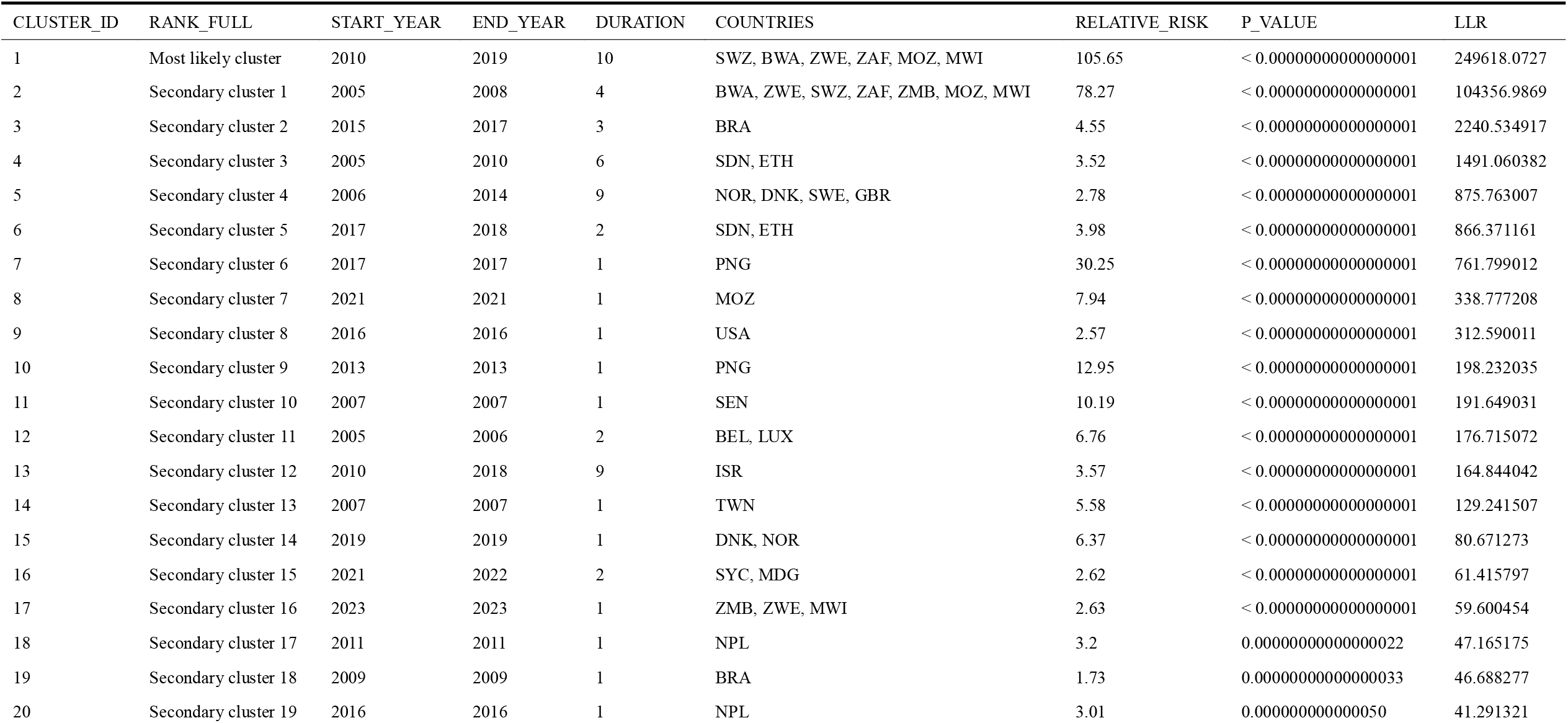
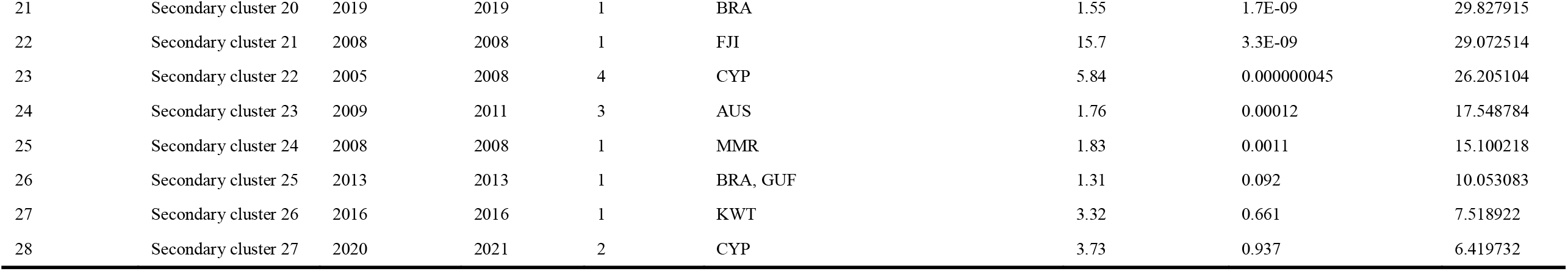
HIV-1 Subtype C Spatiotemporal Clusters - Complete List (2005-2024)

| CLUSTER_ID | RANK_FULL | START_YEAR | END_YEAR | DURATION | COUNTRIES | RELATIVE_RISK | P_VALUE | LLR |
| --- | --- | --- | --- | --- | --- | --- | --- | --- |
| 1 | Most likely cluster | 2010 | 2019 | 10 | SWZ, BWA, ZWE, ZAF, MOZ, MWI | 105.65 | < 0.000000000000000001 | 249618.0727 |
| 2 | Secondary cluster 1 | 2005 | 2008 | 4 | BWA, ZWE, SWZ, ZAF, ZMB, MOZ, MWI | 78.27 | < 0.000000000000000001 | 104356.9869 |
| 3 | Secondary cluster 2 | 2015 | 2017 | 3 | BRA | 4.55 | < 0.000000000000000001 | 2240.534917 |
| 4 | Secondary cluster 3 | 2005 | 2010 | 6 | SDN, ETH | 3.52 | < 0.000000000000000001 | 1491.060382 |
| 5 | Secondary cluster 4 | 2006 | 2014 | 9 | NOR, DNK, SWE, GBR | 2.78 | < 0.000000000000000001 | 875.763007 |
| 6 | Secondary cluster 5 | 2017 | 2018 | 2 | SDN, ETH | 3.98 | < 0.000000000000000001 | 866.371161 |
| 7 | Secondary cluster 6 | 2017 | 2017 | 1 | PNG | 30.25 | < 0.000000000000000001 | 761.799012 |
| 8 | Secondary cluster 7 | 2021 | 2021 | 1 | MOZ | 7.94 | < 0.000000000000000001 | 338.777208 |
| 9 | Secondary cluster 8 | 2016 | 2016 | 1 | USA | 2.57 | < 0.000000000000000001 | 312.590011 |
| 10 | Secondary cluster 9 | 2013 | 2013 | 1 | PNG | 12.95 | < 0.000000000000000001 | 198.232035 |
| 11 | Secondary cluster 10 | 2007 | 2007 | 1 | SEN | 10.19 | < 0.000000000000000001 | 191.649031 |
| 12 | Secondary cluster 11 | 2005 | 2006 | 2 | BEL, LUX | 6.76 | < 0.000000000000000001 | 176.715072 |
| 13 | Secondary cluster 12 | 2010 | 2018 | 9 | ISR | 3.57 | < 0.000000000000000001 | 164.844042 |
| 14 | Secondary cluster 13 | 2007 | 2007 | 1 | TWN | 5.58 | < 0.000000000000000001 | 129.241507 |
| 15 | Secondary cluster 14 | 2019 | 2019 | 1 | DNK, NOR | 6.37 | < 0.000000000000000001 | 80.671273 |
| 16 | Secondary cluster 15 | 2021 | 2022 | 2 | SYC, MDG | 2.62 | < 0.000000000000000001 | 61.415797 |
| 17 | Secondary cluster 16 | 2023 | 2023 | 1 | ZMB, ZWE, MWI | 2.63 | < 0.000000000000000001 | 59.600454 |
| 18 | Secondary cluster 17 | 2011 | 2011 | 1 | NPL | 3.2 | 0.000000000000000022 | 47.165175 |
| 19 | Secondary cluster 18 | 2009 | 2009 | 1 | BRA | 1.73 | 0.000000000000000033 | 46.688277 |
| 20 | Secondary cluster 19 | 2016 | 2016 | 1 | NPL | 3.01 | 0.0000000000000050 | 41.291321 |
| 21 | Secondary cluster 20 | 2019 | 2019 | 1 | BRA | 1.55 | 1.7E-09 | 29.827915 |
| 22 | Secondary cluster 21 | 2008 | 2008 | 1 | FJI | 15.7 | 3.3E-09 | 29.072514 |
| 23 | Secondary cluster 22 | 2005 | 2008 | 4 | CYP | 5.84 | 0.000000045 | 26.205104 |
| 24 | Secondary cluster 23 | 2009 | 2011 | 3 | AUS | 1.76 | 0.00012 | 17.548784 |
| 25 | Secondary cluster 24 | 2008 | 2008 | 1 | MMR | 1.83 | 0.0011 | 15.100218 |
| 26 | Secondary cluster 25 | 2013 | 2013 | 1 | BRA, GUF | 1.31 | 0.092 | 10.053083 |
| 27 | Secondary cluster 26 | 2016 | 2016 | 1 | KWT | 3.32 | 0.661 | 7.518922 |
| 28 | Secondary cluster 27 | 2020 | 2021 | 2 | CYP | 3.73 | 0.937 | 6.419732 |

Table A2 **Error! Reference source not found.** summarizes the country–year hotspot detections stratified by model agreement, listing hotspots identified uniquely by each method and those jointly detected by pairwise or three-method combinations.

**Table A2.**
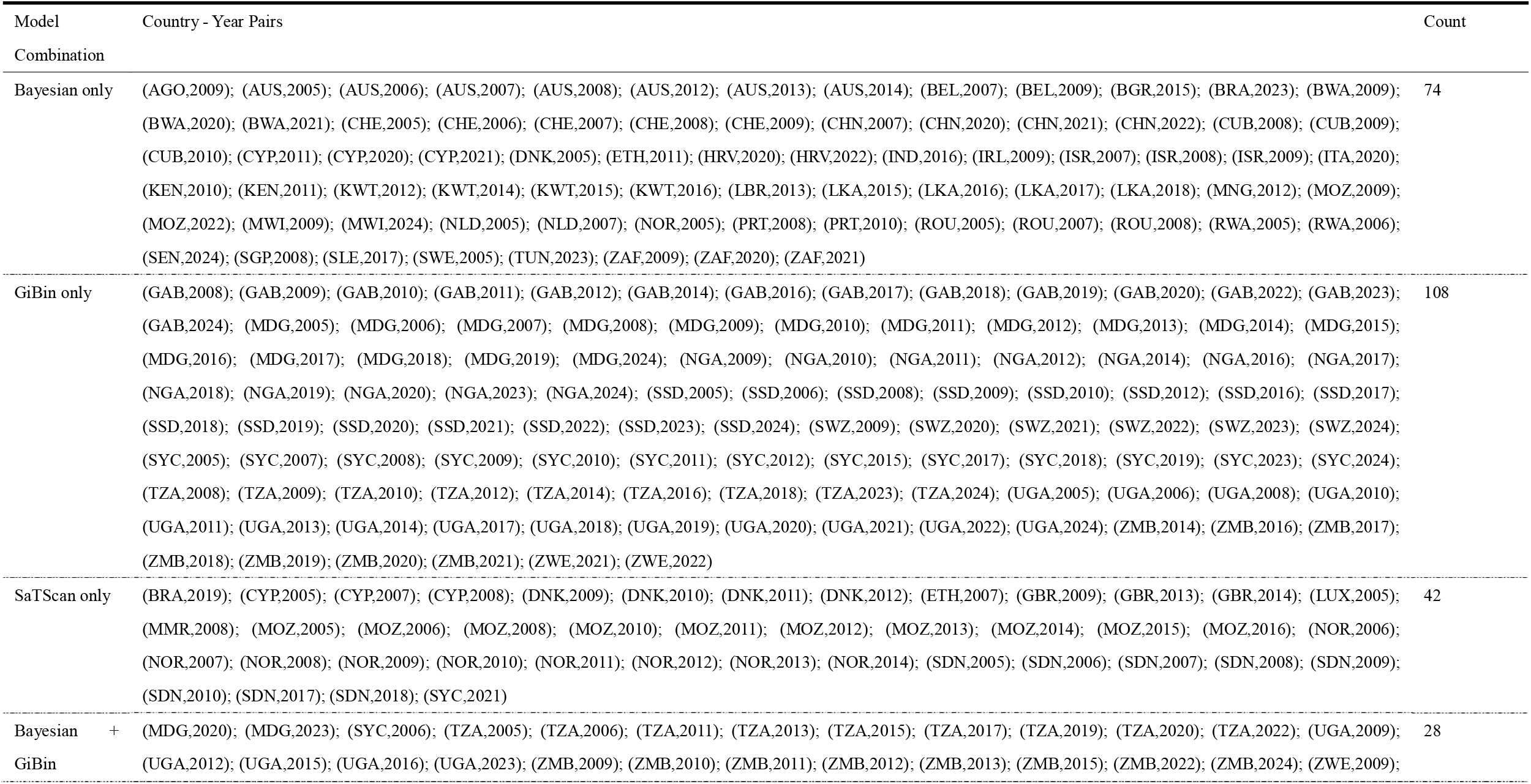
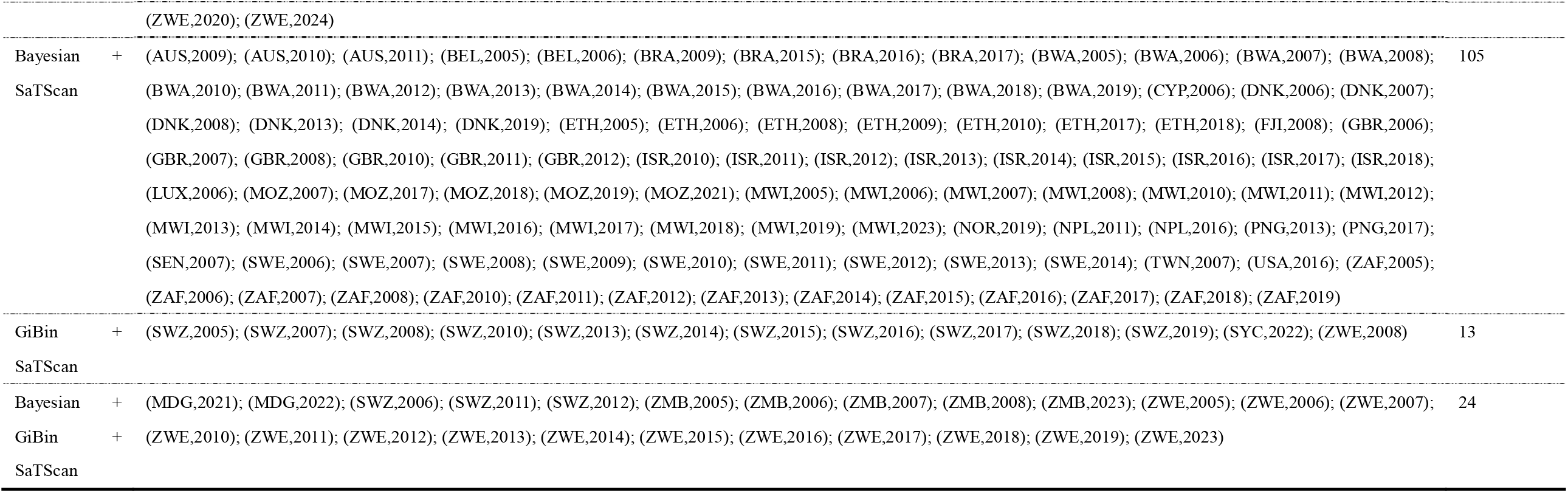
Cross-Model Comparison of Hotspot Detections from Bayesian, GiBin, and SaTScan Methods.

